# RADAR-WMH: Relaxometry And Diffusion Analysis beyond Radiologically defined WMH

**DOI:** 10.64898/2026.07.30.26359145

**Authors:** Vincent Roduit, Fábio Carneiro, Antoine Lutti, Peter Vollenweider, Pedro Marques-Vidal, Julien Vaucher, Martin Preisig, Jean-Philippe Thiran, Aurélie Bussy, Bogdan Draganski

**Affiliations:** Imaging Neuroscience of Ageing laboratory inAGE, Universitätsklinik für Neurologie, Inselspital Bern, University of Bern, Switzerland; Signal Processing Laboratory 5, EPFL-STI-IEL-LTS5, Station 11,CH-1015 Lausanne, Switzerland; Graduate School for Health Sciences, University of Bern, Bern, Switzerland; Centre for Research in Neurosciences, Department of Clinical Neurosciences, Lausanne University Hospital and University of Lausanne, Lausanne, Switzerland; Department of Medicine, Internal medicine, Lausanne University Hospital, Lausanne, Switzerland; Department of Medicine and Specialties, Internal Medicine, Fribourg Hospital and University of Fribourg, Fribourg, Switzerland; Department of Psychiatry, Lausanne University Hospital (CHUV) and University of Lausanne, Prilly, Switzerland; Institute for diagnostic and interventional neuroradiology, Inselspital Bern, University of Bern, Switzerland

**Keywords:** White matter hyperintensities, quantitative MRI, diffusion MRI, machine learning, medical image segmentation

## Abstract

**Background:** White matter hyperintensities (WMH) represent the most visible manifestation of cerebral small vessel disease and of white matter pathology more broadly, yet empirical evidence points to a brain tissue injury extending beyond radiologically detectable lesions on fluid-attenuated inversion recovery (FLAIR) MRI. We present RADAR-WMH (Relaxometry And Diffusion Analysis for Radiological WMH), a multicontrast MRI machine learning framework that characterises white matter pathology through tissue microstructural information rather than lesion contrast alone.

**Methods:** RADAR-WMH was trained on quantitative relaxometry and diffusion-weighted MRI acquired in community-dwelling participants (mean age 59.6 years [SD 22.4], 60.8% women, n=148) using a LightGBM classifier integrating local, textural, and anatomical features at the voxel level. Biological validity was assessed through longitudinal analyses and associations with age, cardio-vascular risk, and cognitive performance in independent cohorts.

**Results:** RADAR-WMH achieved segmentation performance comparable to state-of-the-art FLAIR-based approaches without requiring FLAIR or T1-weighted data. Mean diffusivity was the most influential feature for lesion classification. Beyond FLAIR-defined WMH, RADAR-WMH identified tissue pathology extending outside lesion borders characterised by myelin loss, axonal injury, and increased extracellular water. These microstructural signatures persisted over follow-up and showed stronger association with age, systolic blood pressure, and cognitive performance than corresponding tissue properties restricted to FLAIR-defined WMH extent.

**Conclusions:** RADAR-WMH reveals a significant burden of biologically meaningful white matter injury that remains invisible to FLAIR-defined WMH segmentation. By capturing microstructural pathology linked to vascular risk, cognitive decline, and lesion evolution, RADAR-WMH may provide more sensitive markers of cerebral small vessel disease than FLAIR-visible WMH alone.

## 1. Introduction

White Matter Hyperintensities (WMH) are among the most common incidental findings on brain MRI in older adults, becoming nearly omnipresent beyond age 70 (de Kort et al. 2025; Smith et al. 2017). In the context of ageing, WMH are presumed to reflect cerebral small vessel disease (cSVD) - arteriolosclerotic/vascular risk factor-associated cSVD or cerebral amyloid angiopathy (Duering et al. 2023; Charidimou et al. 2022). WMH are identified as hyperintense lesions on fluid-attenuated inversion recovery (FLAIR) MRI in both clinical and research settings (Duering et al. 2023).

From histological viewpoint, WMH are heterogeneous, showing varying degrees of myelin and axonal loss, astrogliosis, reactive microglia, and markers of blood-brain barrier dysfunction (Wardlaw et al. 2015; Gouw et al. 2011). Critically, these pathological changes are not confined to the FLAIR-visible lesions but extend into the surrounding normal appearing white matter (NAWM) (Solé-Guardia et al. 2025; Fernando et al. 2004; Simpson et al. 2007, 2009), where their presence has been linked to arterial hypertension, underscoring their strong association with cardio-vascular risk (Solé-Guardia et al. 2025).

In vivo characterisation, primarily using diffusion tensor imaging (DTI), has corroborated and extended these histological observations (Bussy et al. 2025). NAWM microstructure is associated with cardio-vascular risk factors including arterial hypertension (Muñoz Maniega et al. 2017; Haight et al. 2018; James et al. 2023; Jochems et al. 2024; Haddad et al. 2022); longitudinal studies demonstrate that NAWM microstructural pathology precedes WMH appearance, identifying regions of heightened vulnerability to lesion progression (van Leijsen et al. 2018; Maillard et al. 2013; Jochems et al. 2024; Maillard et al. 2014-6). There is mounting evidence that NAWM microstructure independently predicts cognitive performance outcome (O’Sullivan et al. 2004; van Norden et al. 2012). More recently, complementary techniques such as Neurite Orientation Dispersion and Density Imaging (NODDI) have further characterized the neurobiology of these changes and their clinical correlates (James et al. 2023; Jochems et al. 2024).

Despite the ample evidence, clinical and research practice continues to rely on FLAIR-defined WMH as the primary measure of white matter disease burden. Because FLAIR provides essentially a binary representation of macrostructurally visible lesions, it inevitably underestimates the true extent of white matter pathology in cSVD. No study to date has attempted to segment white matter lesions based on biophysical modelling-based multi-contrast MRI data. To address this gap, we developed RADAR-WMH - Relaxometry And Diffusion Analysis beyond Radiologically defined WMH, a multicontrast machine learning framework that leverages quantitative relaxometry (MTsat, R1 and R2*) and diffusion-derived metrics (DTI: FA: fractional anisotropy, MD: mean diffusivity; NODDI: ICVF: intracellular volume fraction; ISOVF: isotropic volume fraction; ODI: orientation dispersion index) to identify white matter pathology beyond the binary information brought by FLAIR-defined WMH. We applied RADAR-WMH in the BrainLaus nested subsample of the CoLaus|PsyCoLaus community-based cohort with comprehensive cardio-vascular, cognitive and neuroimaging characterization. Specifically, we aimed to 1) develop a microstructure-based segmentation algorithm independent of FLAIR; 2) compare microstructurally-defined and FLAIR-defined white matter lesion burden; 3) validate the clinical relevance of microstructurally-defined pathology through associations with cardio-vascular risk factors, cognitive outcomes and longitudinal WMH progression.

## 2. Methods

### a. Datasets

Data stem from the BrainLaus (Trofimova et al. 2021), a neuroimaging substudy of CoLaus|PsycoLaus. CoLaus|PsyCoLaus is a longitudinal population-based cohort study of originally 35- to 75-year old participants from the city of Lausanne, Switzerland, designed to assess the associations between mental disorders and cardiovascular risk factors (Firmann et al. 2008). Ethical approval was granted by the Ethics Commission of Canton de Vaud, and participants gave their informed consent prior to participation. The study includes physical and psychiatric baseline evaluations and 3 completed follow-ups. Neuroimaging measures were incorporated from follow-up 2 on, which took place from 2014 to 2018. Four subsamples were included. The first served as training and test set and comprised participants with manually segmented WMH on FLAIR MRI. The remaining three subsamples were used to assess biological validity: 1) a longitudinal subsample to evaluate temporal stability of RADAR-WMH classification, 2) a cross-sectional subsample to examine associations with age, and cognitive outcomes, and 3) a subsample to investigate associations with systolic blood pressure (SBP), one of the main modifiable cardio-vascular risk factors for WMH.

#### i. Model development subsample

The training and test sample was drawn from 163 BrainLaus participants with previously published manual WMH segmentations on FLAIR MRI (Cathala et al. 2025). Participants were excluded for missing MPM or diffusion metrics (n=6) or insufficient image quality on visual assessment (n=9; QC score > 2.5), the latter attributed to motion artifacts, blurring, or processing failures. After exclusion of 15 participants, the final training and test sample comprised 148 individuals. Demographic and biological characteristics, including age, sex, and Fazekas score distribution, are summarized in Table 1.

**Table 1.** Summary of the subsamples and their relevant demographic and clinical characteristics.

Model development subsample
|  | n | Age (mean +/- SD) | Age range (min, max) | Sex (n, %F) | Fazek as 0 (n, %) | Fazek as 1 (n, %) | Fazek as 2 (n, %) | Fazek as 3 (n, %) | Fazek as 4 (n, %) | Fazek as 5 (n, %) | Fazek as 6 (n, %) |
| --- | --- | --- | --- | --- | --- | --- | --- | --- | --- | --- | --- |
| Test | 75 | 58.7 ± 22.4 | 20-93 | 48 (64.0%) | 19 (25.3%) | 10 (13.3%) | 9 (12.0%) | 10 (13.3%) | 9 (12.0%) | 9 (12.0%) | 9 (12.0%) |
| Train | 73 | 60.6 ± 22.4 | 21-93 | 42 (57.5%) | 16 (21.9%) | 9 (12.3%) | 10 (13.7%) | 10 (13.7%) | 10 (13.7%) | 10 (13.7%) | 8 (11.0%) |

Biological validation subsamples
|  | n | Age (mean +/- SD) | Age range (min, max) | Sex (n, %F) | Education (mean years +/- SD) | TMT_A (mean +/- SD) | TMT_B (mean +/- SD) | SBP (mean +/- SD) |
| --- | --- | --- | --- | --- | --- | --- | --- | --- |
| Longitudinal | 12 | 53.9 ± 24.9 | 22.8, 86.2 | 4 (33.3%) | NA | NA | NA | NA |
|  | 12 | 54.9 ± 24.1 | 25.7, 86.3 | 4 (33.3%) | NA | NA | NA | NA |
| Cognition | 286 | 54.0 ± 20.3 | 19.8, 84.2 | 141 (49.3%) | 3.3 ± 1.3 | 33.3 ± 13.5 | 57.5 ± 28.3 | NA |
| SBP | 116 | 72.3 ± 5.7 | 63.3, 84.2 | 60 (51.7%) | 3.1 ± 1.4 | NA | NA | 130.1 ± 18.7 |

#### ii. Biological validation subsamples

Three independent subsamples were used for biological validation, whereas participants involved in model training and testing were excluded from all validation analyses. The longitudinal subsample (n=12) with two MRI timepoints served to evaluate whether RADAR-WMH provides accurate lesion segmentation and captures longitudinal white matter change beyond conventional FLAIR-based detection. A cross-sectional subsample (n=286) with available demographic data and cognitive assessments (Trail Making Test parts A and B), enabled investigation of associations with age and cognitive performance. A subsample of this sample (n=116) with additional SBP data was used to assess sensitivity to cardio-vascular risk. Demographic and biological characteristics of all subsamples are summarized in Table 1.

### b. MRI acquisition

The MRI dataset comprised a FLAIR, quantitative relaxometry, and diffusion-weighted imaging protocols. FLAIR was acquired with TR/TE/TI = 5000/389/1800 ms at 1 mm isotropic resolution. The quantitative relaxometry data was acquired using a custom-made multi-echo 3D FLASH sequence with T1-, PD-, and MT-weighted contrasts (Weiskopf et al. 2013; Draganski et al. 2011). Additionally, B1 mapping data was acquired using a spin-echo/stimulated echo method (Lutti et al. 2010, 2012) to correct for the effect of radio-frequency field inhomogeneities on the relaxometry maps (Lutti et al. 2014). Diffusion-weighted images were acquired using a 2D echo-planar imaging (EPI) sequence at 2 mm isotropic resolution across 118 diffusion directions and multiple b-values (Slater et al. 2019), and B0 field maps for geometric distortion correction. Full acquisition parameters are provided in the Supplementary Methods.

### c. MRI preprocessing

#### i. Quantitative maps

Quantitative maps of magnetization transfer saturation (MTsat), transverse relaxation rate (R2*), effective longitudinal relaxation rate (R1), and effective proton density (PD*) were calculated from the raw MR images using the VBQ toolbox in SPM12 (Draganski et al. 2011; Weiskopf et al. 2013). MTsat and R1 are widely considered markers of myelin content, whereas R2* reflects both myelin and iron content, and PD* primarily reflects the proton density within the voxel (Stüber et al. 2014).

Diffusion MRI preprocessing was performed in MRtrix3 (Tournier et al. 2019), comprising denoising, Gibbs ringing removal (Slater et al. 2019), Eddy current and motion correction (Andersson and Sotiropoulos 2016), and EPI distortion correction using B0 field maps (SPM12 FieldMap toolbox). DTI metrics (FA and MD) were estimated using the tensor model. NODDI parameters (Zhang et al. 2012) (ICVF, ISOVF and OD) were estimated using the AMICO framework (Daducci et al. 2015). MRI g-ratio maps, which represent the ratio between the inner and the outer diameter of the myelin sheath were created.(Stikov et al. 2015; Slater et al. 2019) All maps were rigidly registered to MTsat space and resampled to 1 mm isotropic resolution using FLIRT (Jenkinson et al. 2002). Full preprocessing details are provided in the Supplementary Methods.

#### ii. WMH manual segmentation

Manual WMH segmentation was performed on FLAIR images using the Display tool by a trained rater and reviewed by a neurologist to ensure quality and consistency. The training and test sample was selected to ensure a balanced representation across the full range of Fazekas scores (see Table 1). FLAIR images were linearly registered to MTsat space using FLIRT, and the resulting transformation matrix applied to the manual segmentations using trilinear interpolation.

### d. Model

#### i. Data preparation

Data preprocessing included three steps. First, undefined voxel values (NaN) arising from DTI metrics calculation were set to zero to avoid numerical instabilities. Second, negative voxel values resulting from numerical approximations were truncated to zero. Third, non-brain voxels were removed using a subject-specific brain mask representing the sum of gray matter, white matter and CSF probability maps thresholded at 0.5 in SPM12s “unified segmentation” framework (Ashburner and Friston 2005).

#### ii. Feature selection

In addition to the initial voxel intensity values, we calculated a set of hand-crafted features capturing local context, texture, spatial anatomy, and white matter intensity. These features summarised the local intensity distribution, using the mean and standard deviation within the vicinity of each voxel. Textural features comprised gradient magnitude, Laplacian of Gaussian (LoG), and Local Binary Pattern (LBP), representing edge information and local texture patterns. Spatial features encoded anatomical context, including white matter compartment (periventricular, deep, and superficial white matter) and lobar distribution (frontal, parietal, temporal, and occipital) derived from SPM12s neuromorphometric atlas. Finally, a White Matter Intensity Energy (WMIE) feature was introduced to enhance the discrimination between NAWM and WMH. A detailed mathematical description of all features is provided in the Supplementary Methods.

#### iii. Light gradient boost

LightGBM (LGBM), a tree-based algorithm, was used for classification. As an ensemble method, LGBM builds trees sequentially, enabling the modeling of complex non-linear relationships. Its histogram-based split finding and efficient training strategy allow for faster training and lower memory usage than alternative implementations such as XGBoost, making it well suited to our dataset size. Hyperparameter optimisation ranges are summarized in Supplementary Table 1.

**Figure 1:**
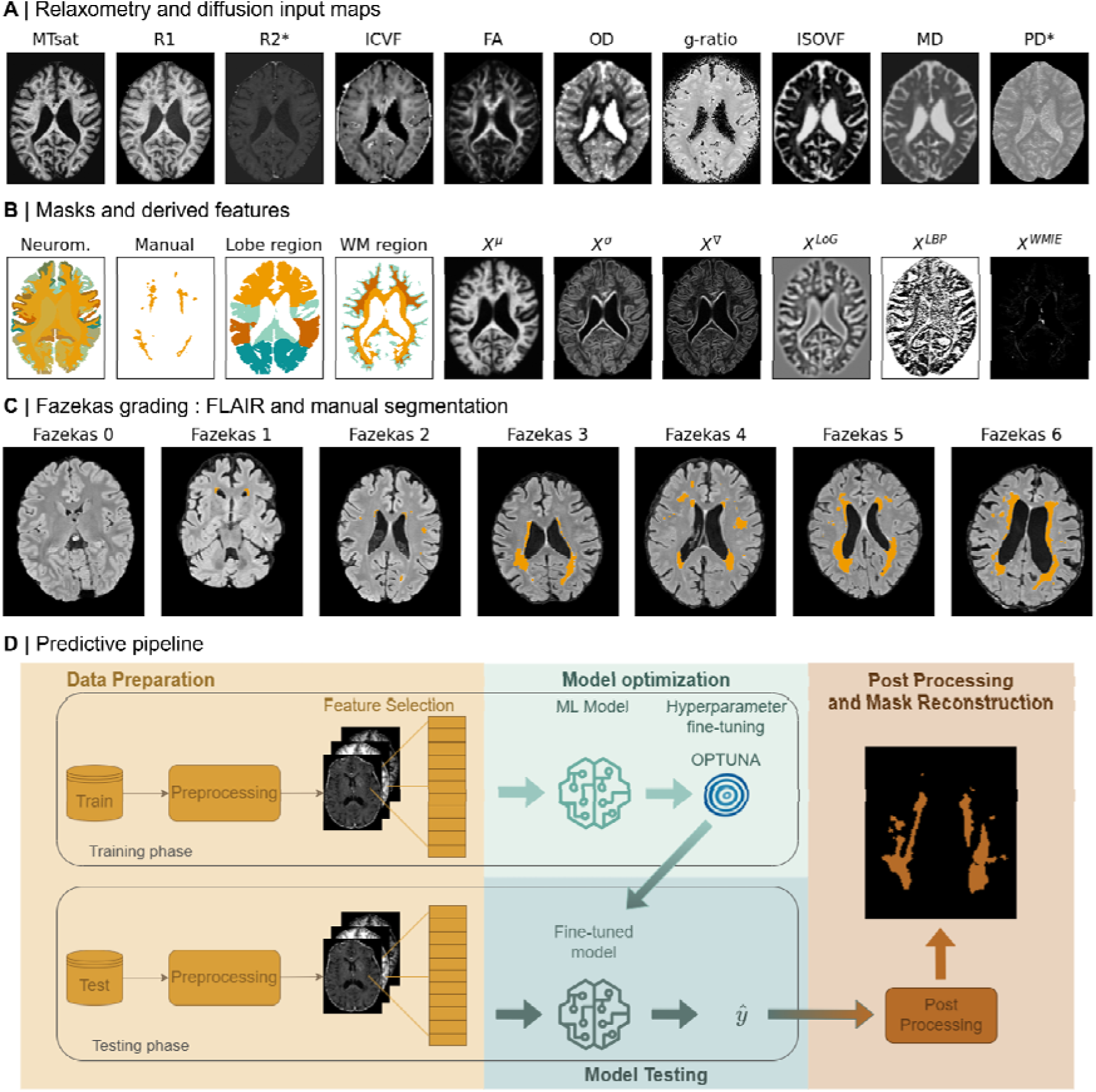
**A)** Representative dataset example showing myelin- and iron-sensitive metrics (magnetization transfer saturation [MTsat], longitudinal relaxation [R1], transverse relaxation [R2*]), axon-sensitive metrics (intracellular volume fraction [ICVF], fractional anisotropy [FA], orientation dispersion [OD], g-ratio), and fluid metrics (isotropic volume fraction [ISOVF], mean diffusivity [MD]). **B)** Neuromorphometric mask, manual segmentation, and lobe/white matter (WM) region masks, with examples of features derived from an MTsat map: local information (mean, standard deviation), textural information (gradient, Laplacian of Gaussian, Local Binary Pattern, and White Matter Intensity Energy). **C)** Illustrative Fazekas scores ranging from 0 (no WMH) to 6 (severe deep and periventricular WMH). **D)** Predictive pipeline. The training phase includes data preparation and model training. The testing phase applies the same preprocessing, followed by model inference and post-processing for mask reconstruction.

#### iv. Model assessment

Multiple model variants were evaluated by systematically combining input features and preprocessing pipelines, with all models trained, validated, and optimised under identical conditions. The hyperparameter search space is shown in Figure 2A, and optimization was performed using Optuna (Akiba et al. 2019).

**Figure 2:**
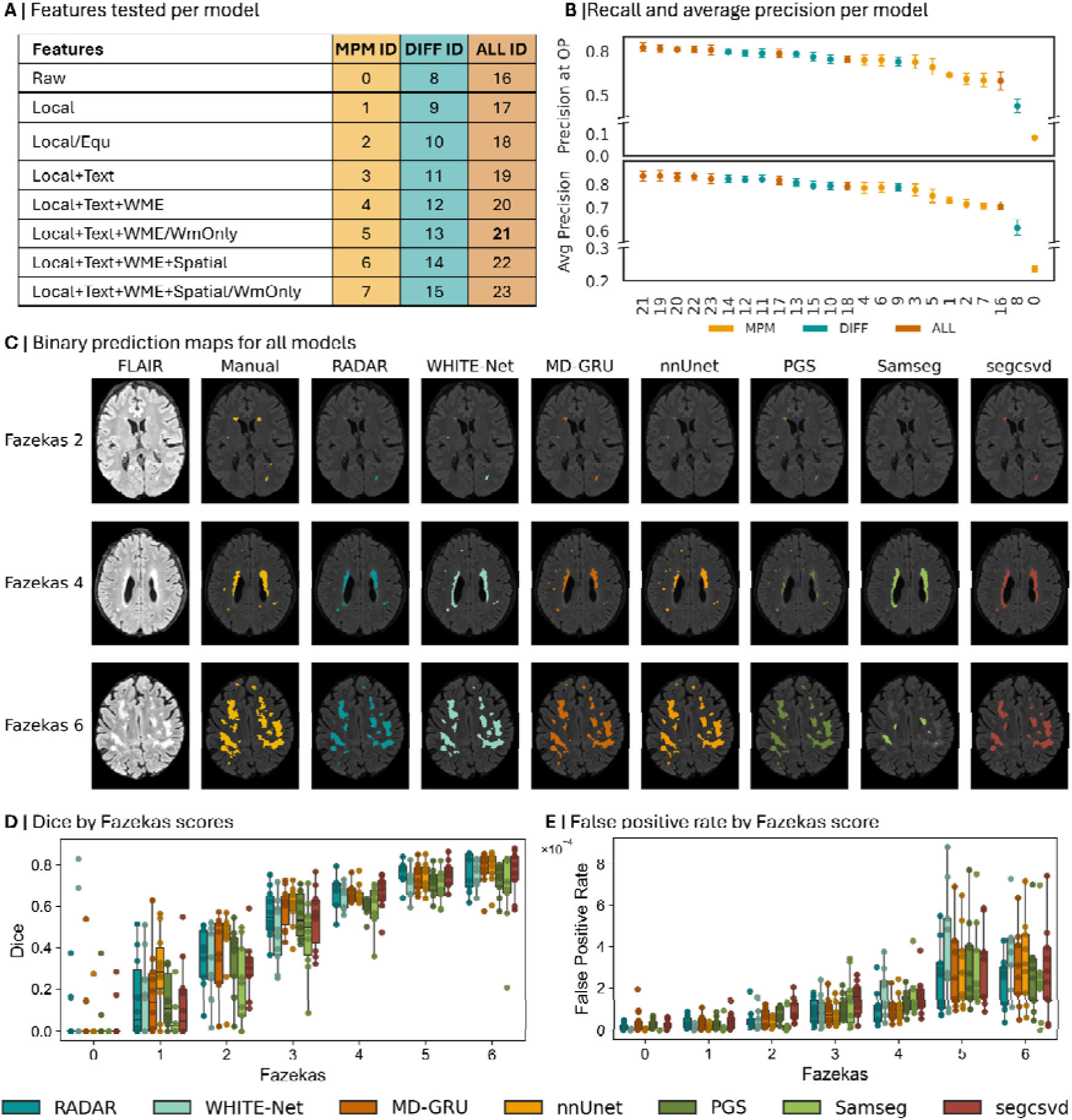
**A)** Model IDs organized by feature set and modality. **B)** Precision at an operating point (OP) with recall = 0.7, along with average precision across all models. **C)** Axial view of FLAIR images, manual segmentations, and binary prediction maps for all models in three example cases: Fazekas scores 2, 4, and 6; sagittal view in Supplementary figure 2. **D)** Dice score per method across Fazekas scores (0-6). **E)** False positive rate (FPR) per method across Fazekas scores (0-6).

The analysis comprised three components: (i) comparison of MRI contrasts and derived maps (diffusion-only, relaxometry-only, and combined), motivated by the limited availability of open access relaxometry MRI data; (ii) comparison of preprocessing strategies (Section 2.d.ii); and (iii) their combined impact on lesion segmentation performance.

Performance was assessed using voxel-wise precision-recall (PR) curves with five-fold cross-validation. An operating point (OP) was defined at a fixed recall of 0.7, and median average precision (AP) with 95% confidence intervals (CI) was reported. Metrics were calculated without post-processing to allow fair comparison of raw model outputs. Training was restricted to participants with Fazekas scores >3, and to a subset of 10 voxels for computational efficiency, as inclusion of all voxels yielded only marginal performance gains.

#### v. Post-processing

In a post-processing step, we aimed to reduce false positives and improve segmentation quality. First, we enforced a minimum lesion size threshold of five voxels, removing small connected components defined using 18-connectivity to avoid splitting biologically related lesions. Second, we applied anatomical constraints using a WM mask dilated by two voxels to account for boundary inaccuracies. Lesions with more than 50% of voxels outside this mask were classified as false positives and removed. This approach reduced false detections while preserving true positives.

#### vi. Model interpretability

Model interpretability remains a key challenge in machine learning, as many models operate as “black boxes” with limited insight into their predictions (Hassija et al. 2024). Here, interpretability was assessed using SHapley Additive exPlanations (SHAP) (Mitchell et al. 2022; Lundberg and Lee 2017), which quantify each feature’s contribution relative to a baseline model output. For LGBM, this corresponds to the average prediction in log-odds. SHAP values indicate how individual features increase or decrease the probability of a voxel being classified as WMH, enabling interpretation of the model’s decision process and assessment of the biological plausibility of learned patterns. Further methodological details are provided in the Supplementary Methods.

### e. Model comparisons

The best-performing LightGBM model was compared against existing WMH segmentation methods. As no models trained on MPM- or diffusion-derived metrics are currently available, comparisons were limited to approaches based on FLAIR and/or T1-weighted images. These included U-Net–based models (WHITE-Net (Cathala et al. 2025), nnU-Net (Gesierich et al. 2025), segcsvd (Gibson et al. 2026), PGS (Park et al. 2021)), as well as MD-GRU (Gesierich et al. 2025) and Samseg (Puonti et al. 2016). WHITE-Net was developed on FLAIR-only images acquired from the same BrainLaus subjects, while RADAR-WMH was trained on non-FLAIR multi-contrast MRI data.

Performance was evaluated using standard segmentation metrics, including Dice Similarity Coefficient (DSC), precision, recall, volume similarity (VS), average symmetric surface distance (ASSD), relative volume difference (RVD), 95th percentile Hausdorff distance (HD95), and weighted Dice coefficient (WDC). Detailed definitions are provided in the Supplementary Methods. Given dataset heterogeneity, metric estimates were obtained via bootstrapping and reported as mean values with corresponding 95% confidence intervals (Maier-Hein et al. 2018).

### f. Uncertainty quantification

Beyond producing a binary segmentation, reliable interpretation of model predictions requires quantifying the confidence that can be attributed to them. Uncertainty was therefore evaluated using Shannon Entropy (H) at three complementary scales - voxel-, lesion-, and participant-level, each capturing different types of potential errors. Equations are provided in the Supplementary Methods.

### g. Model biological sensitivity

These analyses evaluated the added value and potential limitations of a microstructure-based WMH segmentation approach (RADAR-WMH) relative to a FLAIR-based method. WHITE-Net tool served as the reference comparator, having been trained on the same sample and demonstrated performance comparable to RADAR-WMH, particularly in Dice similarity. Specifically, we assessed whether the two approaches provide concordant information on longitudinal WMH change and on sensitivity to key risk factors such as age and SBP, and compared their ability to predict cognitive performance.

#### i. Longitudinal assessment

Twelve participants with two FLAIR MRI timepoints were included. For each participant, FLAIR, MPM, and diffusion-derived maps were co-registered to a common subject-specific space, and binary WMH masks were generated at each timepoint using WHITE-Net (FLAIR) and RADAR-WMH (multi-contrast MRI). Lesions were defined as 3D connected components using 18-connectivity with a minimum size of 5 voxels, and lesion correspondence across methods and timepoints was established spatially via overlap of connected components on a common voxel grid.

At the second timepoint, WHITE-Net lesions were classified as new, extending, regressing, or stable based on overlap with baseline lesions and relative volume change (>10%, <-10%, or within ±10%). At the voxel level, lesions were similarly categorised as emerging, disappearing or stable. For example, RADAR-WMH-only lesions were classified according to whether they subsequently became visible only on WHITE-Net, on both methods, remained RADAR-WMH-only, or disappeared.

Group differences in lesion probability, uncertainty, and regional quantitative MRI measures were assessed using Mann-Whitney U or paired Wilcoxon signed-rank tests, with Benjamini-Hochberg false discovery rate correction for multiple comparisons. Further details on the processing and statistical analyses are provided in the Supplementary Methods.

#### ii. Age, cardio-vascular risk factor and cognition

Biological validation analyses compared the sensitivity of RADAR-WMH- and WHITE-Net-derived WMH metrics to clinically relevant variables, including age, SBP, and cognitive performance. Here, we used measurements of processing speed with the Trail-Making test (TMT) Part A and executive function Part B (TMT-B). Additional description of the tests can be found in Supplementary methods. WMH volume and quantitative MRI metrics extracted within WMH were compared between methods, and their associations with clinical variables were assessed using paired regression analyses accounting for within-subject measurements. The incremental predictive value of each MRI metric beyond demographic covariates was further evaluated using repeated cross-validated ridge regression. Statistical significance was assessed using FDR correction and, where appropriate, permutation testing. Full methodological details are provided in the Supplementary Methods.

## 3. Results

### a. Model selection

Segmentation results obtained for all the models (Figure 2A) are shown in Figure 2B. Model ID 21 achieved the highest recall at the OP (0.826 [0.775, 0.857]), while Model ID 19 attained the highest AP (0.838 [0.816, 0.861]). Considering the trade-off between AP and OP, Model ID 21 was selected for subsequent analyses. This model combines local, textural, and WMIE features and applies WM-based restriction during preprocessing.

Overall, models based solely on relaxometry MRI showed the lowest performance, followed by diffusion-only models, while combining both modalities yielded the best results. Performance consistently improved with the inclusion of additional features and preprocessing steps compared to raw MRI inputs, with local features contributing most to this gain. Notably, diffusion-only models remained competitive (e.g., ID 14: AP 0.825 [0.811, 0.840], OP 0.797 [0.781, 0.814]), indicating that diffusion metrics are the primary drivers of performance, with relaxometry providing only modest additional benefit.

### b. Model comparison

Table 2 summarizes the results across all evaluated methods. Samseg achieved the highest performance on several metrics, including DSC (0.56 [0.48, 0.64]), volume similarity (VS: 0.71 [0.62, 0.79]), ASSD (4.24 [2.62, 6.29]), and WDC (0.57 [0.49, 0.65]). nnU-Net performed best on sensitivity, RVD, HD95, and WDC, while the highest sensitivity was also observed for MD-GRU and segcsvd. Although the proposed RADAR-WMH model did not achieve the top performance, it remained competitive (DSC: 0.41 [0.34, 0.48]), with results comparable to WHITE-Net (DSC: 0.42 [0.35, 0.49]), which was trained on the same dataset.

**Table 2:** Performance comparison across models. The mean value across participants with 95% bootstrap CI is presented. The best value per metric is highlighted in **bold**. Dice-Sørensen Coefficient (DSC), volume similarity (VS), average symmetric surface distance (ASSD), relative volume difference (RVD), 95 Hausdorff Distance (HD95), weighted Dice coefficient (WDC).

| Model | DSC | Precision | Recall | VS | ASSD | RVD | HD95 | WDC |
| --- | --- | --- | --- | --- | --- | --- | --- | --- |
| WHITE-Net | 0.42<br>[0.35, 0.49] | 0.45 [0.37, 0.52] | 0.44<br>[0.36, 0.51] | 0.60<br>[0.52, 0.69] | 7.32<br>[3.98, 11.39] | 0.53<br>[0.30, 0.85] | 17.43<br>[12.08, 23.35] | 0.45<br>[0.38, 0.52] |
| MD-GRU | 0.42<br>[0.35, 0.49] | 0.45 [0.36, 0.52] | **0.46**<br>**[0.39, 0.52]** | 0.58<br>[0.49, 0.67] | 7.52<br>[4.22, 11.39] | 3.54<br>[0.72, 7.90] | 16.81<br>[11.04, 23.38] | 0.47<br>[0.40, 0.55] |
| nnUnet | 0.49<br>[0.42, 0.56] | **0.47**<br>**[0.39, 0.55]** | 0.44<br>[0.38, 0.51] | 0.67<br>[0.59, 0.75] | 4.93<br>[1.84, 8.99] | **0.52**<br>**[0.25, 0.90]** | **10.84**<br>**[6.64, 15.95]** | 0.56<br>[0.49, 0.63] |
| segcsvd | 0.40<br>[0.33, 0.47] | 0.41 [0.33, 0.49] | **0.46**<br>**[0.39, 0.53]** | 0.54<br>[0.45, 0.63] | 7.32<br>[4.33, 10.80] | 2.10<br>[1.04, 3.45] | 17.84<br>[11.78, 24.62] | 0.43<br>[0.36, 0.51] |
| PGS | 0.38<br>[0.32, 0.45] | 0.41 [0.33, 0.48] | 0.45<br>[0.38, 0.51] | 0.54<br>[0.46, 0.63] | 7.14<br>[4.10, 10.73] | 1.89<br>[0.93, 3.07] | 17.88<br>[11.83, 24.83] | 0.44<br>[0.37, 0.52] |
| Shiva | 0.37<br>[0.31, 0.44] | **0.47**<br>**[0.39, 0.56]** | 0.39<br>[0.32, 0.46] | 0.52<br>[0.44, 0.60] | 6.85<br>[3.82, 10.58] | 1.72<br>[0.51, 3.75] | 17.57<br>[11.71, 24.21] | 0.46<br>[0.39, 0.52] |
| Samseg | **0.56**<br>**[0.48, 0.64]** | 0.40 [0.31, 0.48] | 0.32<br>[0.26, 0.39] | **0.71**<br>**[0.62, 0.79]** | **4.24**<br>**[2.62, 6.29]** | 0.89<br>[0.35, 1.77] | 13.53<br>[10.04, 17.32] | **0.57**<br>**[0.49, 0.65]** |
| RADAR | 0.41<br>[0.34, 0.48] | 0.44 [0.36, 0.52] | 0.43<br>[0.36, 0.50] | 0.58<br>[0.49, 0.66] | 7.46<br>[4.33, 11.06] | 1.87<br>[0.70, 3.42] | 18.54<br>[12.61, 25.19] | 0.44<br>[0.36, 0.51] |

Qualitative examples (Figure 2C) show broadly similar segmentation outputs across methods, with lower performance observed for Samseg in high lesion load cases (Fazekas 6). Quantitative analysis across Fazekas groups (Figure 2D-E) shows that DSC improves with increasing lesion burden, while the false positive rate also increases. Samseg shows particularly high DSC performance in Fazekas 0 cases (DSC: 0.84 [0.68, 1.00]), which strongly influences overall results given their proportion in the dataset (25%). When excluding Fazekas 0 cases (Supplementary table 2), performance rankings change substantially: nnU-Net achieves the best overall performance (DSC: 0.58 [0.52, 0.63]), while Samseg becomes the lowest-performing method (DSC: 0.46 [0.38, 0.53]). The proposed model achieves a DSC of 0.54 [0.47, 0.60], comparable to WHITE-Net, MD-GRU, and segcsvd. Overall, the proposed RADAR-WMH method achieves competitive performance relative to state-of-the-art approaches, and demonstrates the overall lower false positive rate (FPR: 9.97×10^-5^ (7.26×10^-5^, 1.30×10^-4^) despite not relying on FLAIR or T1-weighted imaging.

### c. Model interpretation

#### i. Feature importance

Feature importance was analyzed using SHAP values (Figure 3A), with features ranked by mean absolute contribution. To ensure reliable interpretation, only true positive (TP) and true negative (TN) voxels were considered (n = 47,163). Mean diffusivity (MD) emerged as the most influential feature. Notably, six of the nine most important features (MD mean, ODmean, R2* mean, g-ratio std, ICVF mean, ISOVF mean) were local features, highlighting their strong predictive value. Additionally, edge-related features (MD LoG, MTsat LoG) contributed to lesion discrimination, indicating the importance of boundary information.

**Figure 3.**
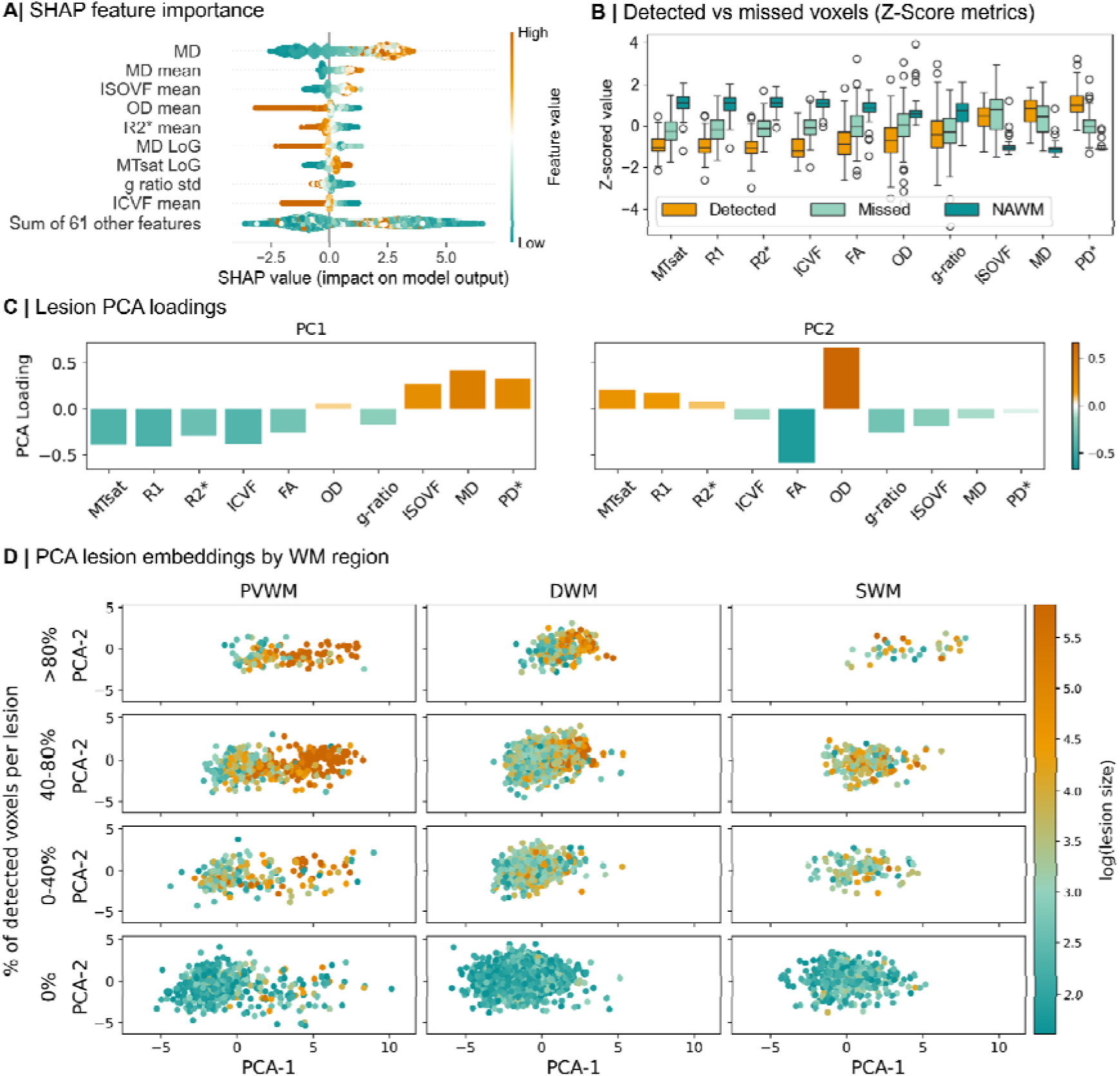
**A)** SHAP beeswarm plot for the LGBM model, with features ranked by mean absolute SHAP value; SHAP values are shown on the x-axis and feature values are color-coded (blue: low, orange: high). **B)** Z-scored differences across metrics between detected voxels (orange), missed voxels (light blue) and NAWM reference voxels (dark blue). **C)** PCA loadings of lesion features, where the first two components (PC1 and PC2) explain 45% and 18% of the variance, respectively; colors indicate loading strength (blue: negative, orange: positive). **D)** PCA embedding of lesions grouped by white matter regions (SWM, DWM, PVWM), with color representing log-transformed lesion size (blue: small, orange: large).

SHAP values also provide insight into feature directionality. Higher MD values (orange) increased the probability of WMH classification (positive SHAP value), consistent with increased extracellular water content in lesions, while higher ICVF values, reflecting axonal integrity, reduced this probability. Overall, six of the nine most important features were diffusion-derived, supporting earlier findings that diffusion metrics drive model performance, with relaxometry features providing only limited additional benefit.

#### ii. Segmentation difficulty

This analysis investigates factors contributing to segmentation errors by comparing MRI metric properties of detected and missed voxels. For each participant, differences between the two groups were assessed using the Wilcoxon signed-rank test, excluding lesions smaller than five voxels. All metrics showed significant differences (FDR-corrected, p < 0.05, Supplementary Table 3), with medium to large effect sizes, except for g-ratio, MD, and ISOVF, which showed smaller effects. To visualize these differences, MRI metrics were z-scored (Figure 3B). Detected voxels exhibited pronounced microstructural alterations compared to NAWM (Supplementary Table 3), including reduced myelin-related measures, increased free water content, and greater axonal disruption. In contrast, missed voxels showed significantly less severe abnormalities than detected voxels, suggesting that these voxels are missed because they reflect relatively healthier tissue.

To further characterize differences between detected and missed lesions, a principal component analysis (PCA) was performed. Lesions were grouped based on the proportion of correctly detected voxels (0%, 0-40%, 40-80%, >80%). Figure 3C demonstrated that PC1 was mainly driven by negative MTsat, R1, R2*, ICVF, and positive MD, ISOVF, PD*, while PC2 was driven by positive OD and negative FA. In Figure 3D, The PCA projection showed that lesions with higher detection rates were generally larger and exhibited higher PC1 values, reflecting more pronounced microstructural impairment (e.g., reduced myelin, increased edema, and axonal damage). In contrast, smaller lesions showed lower PC1 values and were less frequently detected, suggesting that less severely affected tissue is more difficult to segment. Overall, these findings indicate that the degree of microstructural impairment is related to the lesion size, and that both are key factors driving segmentation performance.

### d. Model uncertainty

Model uncertainty at three spatial scales is illustrated in Figure 4A. At the voxel level, uncertainty is highest at lesion boundaries, indicating that lesion borders are the most challenging regions to predict. In contrast, uncertainty within lesion cores is low, reflecting high model confidence in these areas. At the lesion level, larger lesions exhibit lower uncertainty, whereas smaller lesions are associated with higher uncertainty. At the participant level, uncertainty provides a global estimate of segmentation reliability, with individuals exhibiting lower lesion burden showing higher overall uncertainty. Figure 4B shows that participant-level uncertainty is negatively correlated with the Dice score, indicating that lower segmentation quality is associated with higher uncertainty. However, as shown in Figure 4C, this relationship is partly driven by lesion burden, as the correlation is reduced after residualizing for Fazekas scores. This suggests that both Dice score and uncertainty are similarly influenced by overall lesion load.

**Figure 4:**
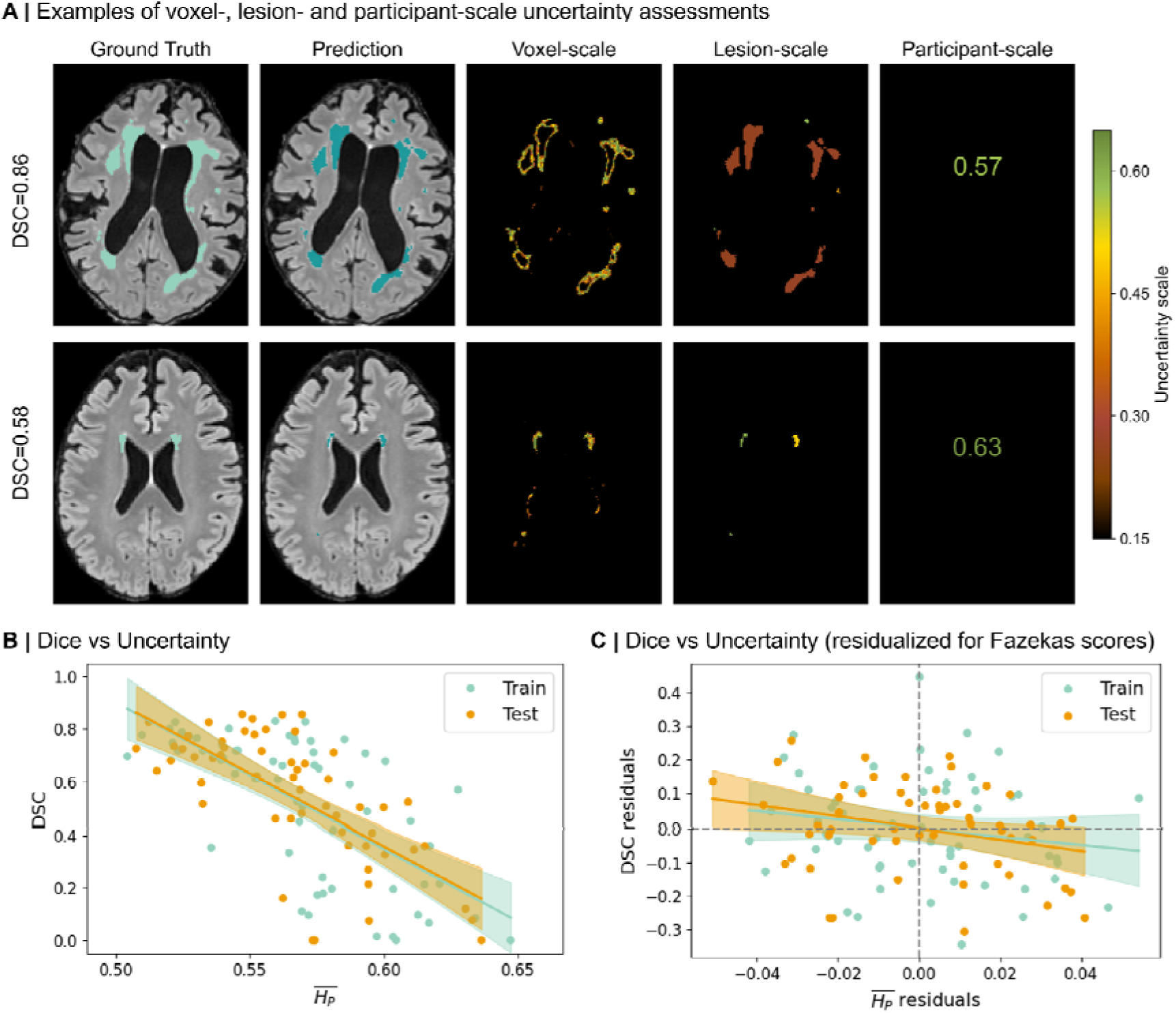
**A)** Axial views of uncertainty maps at multiple scales. The first column shows manual segmentations overlaid on FLAIR images, the second column shows predictions from the best-performing model, and the last three columns display uncertainty at the voxel-, lesion-, and participant-level, respectively (sagittal view in Supplementary figure 3). **B)** Scatter plot of training and test data, showing a negative association between_and Dice score (DSC). **C)** Residual scatter plot of_versus DSC after adjusting for Fazekas score, demonstrating an attenuated correlation after accounting for lesion burden.

### e. Longitudinal analysis

Figure 5 summarizes the longitudinal behavior and qMRI characteristics of segmentation-specific lesion voxels identified by RADAR-WMH and WHITE-Net. RADAR-WMH identified significantly more new lesions (median [25-75 IQR]: RADAR-WMH 4 [1.5-8.8], WHITE-Net 2 [0-6]), whereas the numbers of extending, stable, and regressing lesions were comparable between methods (Figure 5A). Lesion volumes showed comparable distributions between methods except in the regressing lesions showing lower volume in WHITE-Net (20 [0-304]) vs RADAR (203 [111-495.5]) but differed substantially across lesion categories, with extending, stable and regressing lesions exhibiting the largest volumes and new lesions being consistently smaller (Figure 5B). Within each method, longitudinal overlap between timepoint 1 (T1) and timepoint 2 (T2) segmentations remained moderate to high (Figure 5C), suggesting that the segmentations were relatively stable across time despite expected biological lesion evolution. In contrast, overlap between the two methods computed at the same timepoint was substantially lower (Figure 5D), indicating systematic differences in the lesions identified by each method.

**Figure 5.**
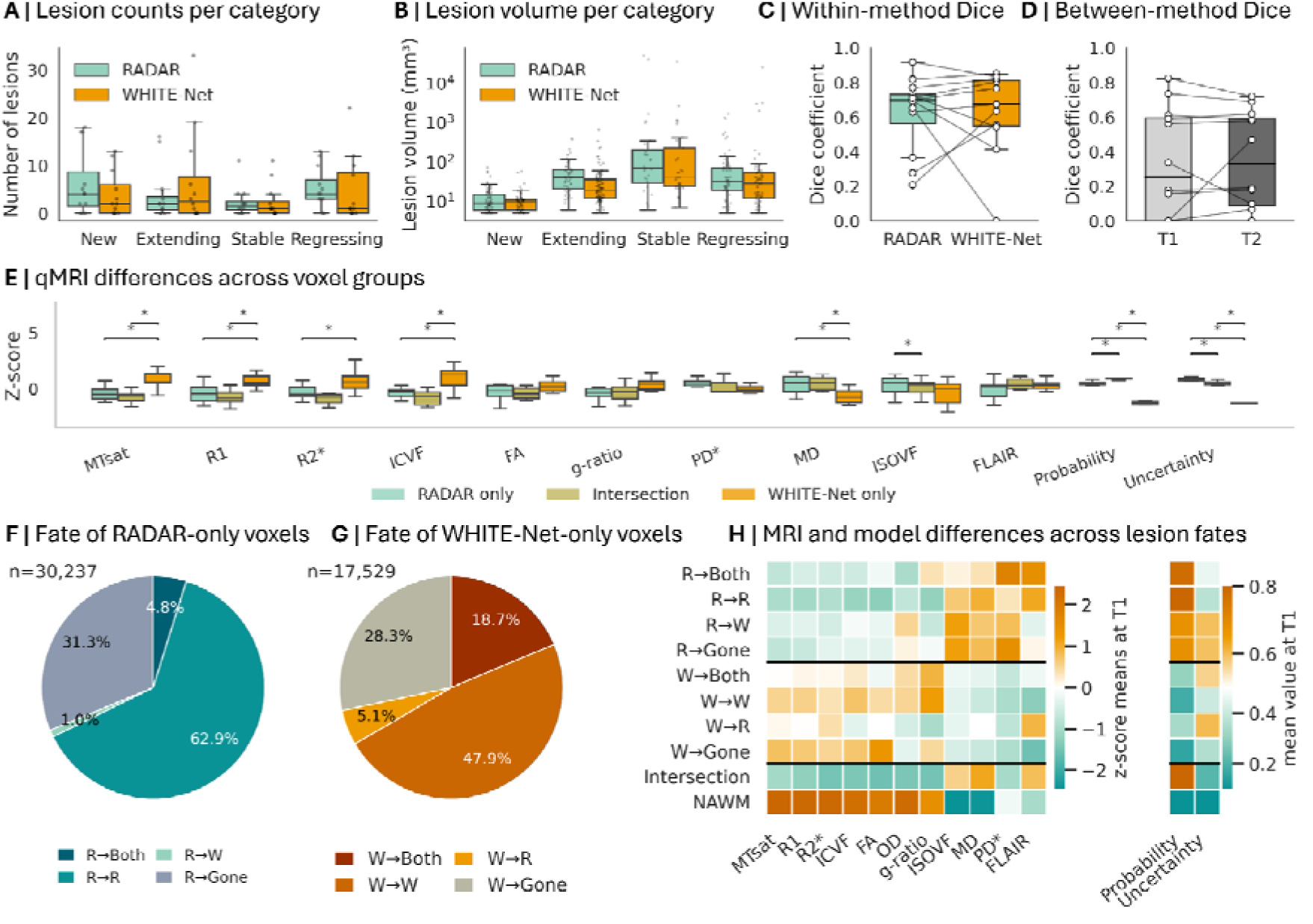
Longitudinal evolution and qMRI characteristics of segmentation-specific white matter lesion voxels identified by RADAR-WMH and WHITE-Net. **(A)** Distribution of the number of lesions per participant classified as new, extending, stable, or regressing, stratified by method. **(B)** Distribution of lesion volumes across lesion categories (log scale). Boxes indicate median and interquartile range. **(C)** Within-method longitudinal spatial overlap (Dice coefficient) between timepoint 1 (T1) and timepoint 2 (T2) segmentations. **(D)** Between-method spatial overlap between RADAR-WMH and WHITE-Net segmentations at T1 and T2. **(E)** qMRI differences across segmentation-specific voxel groups (RADAR-WMH only, Union, and WHITE-Net only). Values are shown as z-scores relative to normal-appearing white matter. Asterisks indicate significant pairwise differences after multiple-comparison correction. **(F)** Longitudinal fate of RADAR-only voxels identified at T1. (G) Longitudinal fate of WHITE-Net-only voxels identified at T1. **(H)** Mean qMRI z-scores across longitudinal voxel fate categories at T1. Warmer colors indicate higher z-scores, and cooler colors indicate lower z-scores relative to normal-appearing white matter. **Abbreviations: T1**, Timepoint 1; **T2**, Timepoint 2. R→Both, lesions segmented by RADAR only at T1 and by both methods at T2; R→R, segmented by RADAR only at both T1 and T2; R→W, segmented by RADAR only at T1 and by WHITE-Net only at T2; R→Gone, segmented by RADAR only at T1 and not detected by either method at T2. The same notation applies to lesions initially segmented by WHITE-Net only: W→Both, W→R, W→W, and W→Gone.

To better understand the tissue characteristics captured by these method-specific segmentations, we next investigated the microstructural properties of voxels belonging to the intersection of both methods (i.e., voxels identified by both methods) versus voxels uniquely identified by each method. Segmentation-specific voxel groups exhibited distinct qMRI profiles (Figure 5E). Compared with intersection voxels, RADAR-only voxels showed similar values across most metrics, except for higher ISOVF, lower probability and higher uncertainty. In contrast, WHITE-Net-only voxels showed higher MTsat, R1, and ICVF values, together with lower MD, lesion probability and uncertainty. These findings suggest that RADAR-only voxels may capture tissue with microstructural abnormalities more comparable to voxels identified by both methods, while additionally detecting voxels with greater free water content despite higher uncertainty. In contrast, WHITE-Net-only voxels may preferentially identify tissue with relatively preserved microstructural integrity.

Longitudinal fate analysis showed that most RADAR-only or WHITE-Net-only voxels at T1 either became identified by both methods at T2 or remained method-specific (Figure 5F-G). Transitions from one method-specific category to the opposite method were comparatively uncommon for both approaches. RADAR-only voxels at T1 that remained RADAR-specific or identified by both methods at T2 showed the strongest qMRI evidence of microstructural damage, with lower MTsat, R1, ICVF, FA, OD, and g-ratio, alongside higher ISOVF, MD, PD*, FLAIR intensity, lesion probability, and lower uncertainty - a profile similar to the microstructural pattern of the intersection voxels (Figure 5H). By contrast, RADAR-only voxels at T1 that either disappeared at T2 or were subsequently detected only by WHITE-Net exhibited a similar pattern of microstructural abnormalities, despite substantially lower FLAIR intensity. This finding suggests that FLAIR signal intensity is relatively insensitive to the underlying microstructural damage of these voxels and may explain their omission by WHITE-Net. In contrast, WHITE-Net-only voxels at T1 exhibited less pronounced qMRI abnormalities, irrespective of whether they disappeared or persisted at T2. Their microstructural profile more closely resembled that of NAWM, characterized by relatively preserved myelin- and axonal-related metrics and lower water accumulation. In addition, they also showed lower lesion probability, higher uncertainty, and lower FLAIR intensity.

Only voxels later detected by RADAR-WMH at T2 showed high FLAIR hyperintensity, suggesting that most WHITE-Net-only voxels do not exhibit clear hyperintense FLAIR signal and instead correspond to tissue with comparatively preserved microstructural properties.

### f. Biological validation

In the cognitive sample (n=286), we compared the overall distributions of WMH volume and microstructural measures of the WMH defined by the two methods (Figure 6A). Significant differences were observed, with RADAR-WMH yielding larger WMH volumes together with more abnormal tissue properties, characterized by lower R2*, ICVF, FA, and g-ratio values, and higher MD and PD* values compared with WHITE-Net.

**Figure 6:**
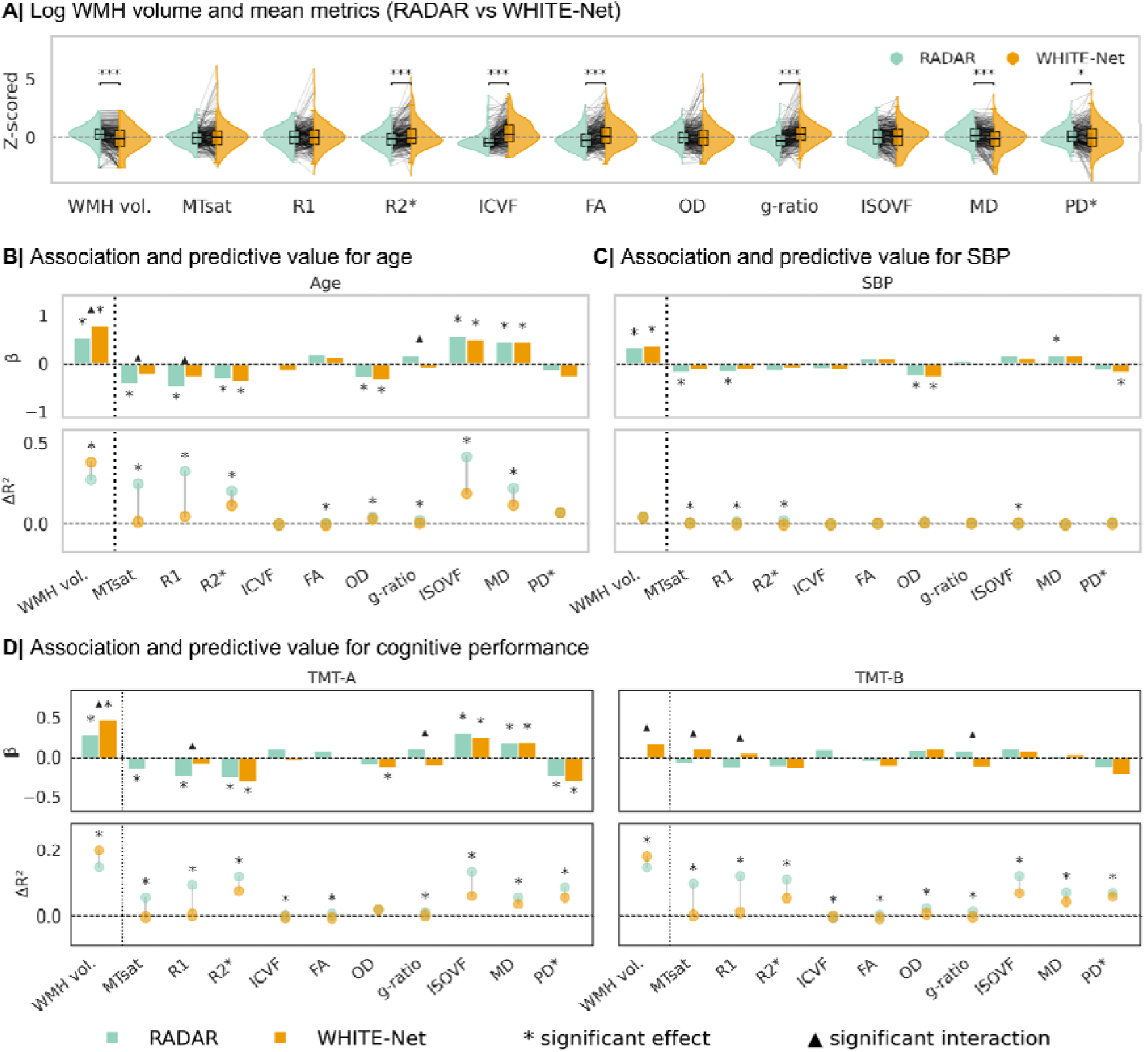
Comparison of RADAR- and WHITE-Net-derived MRI metrics and their association with age, systolic blood pressure, and cognitive performance. (**A**) Distribution of log-transformed WMH volume and mean microstructural metrics extracted within WMH from RADAR-WMH and WHITE-Net segmentations. Metrics were jointly z-scored within each metric across methods. Half-violin plots, paired subject-level trajectories, and boxplots are shown. Statistical significance was assessed using paired t-tests with FDR correction. Association and predictive value analyses for age (**B**), systolic blood pressure (SBP; **C**), and cognitive performance measured using Trail Making Test A and B (TMT-A and TMT-B; **D**). Upper panels show standardized regression coefficients (β) from paired linear models relating MRI metrics to each clinical variable. Lower panels show the additional cross-validated predictive value (ΔR²) obtained when adding each MRI metric to a baseline model including sex and education, reflecting variance explained beyond covariates alone. ΔR² values from RADAR- and WHITE-Net-derived metrics were directly compared using permutation testing. Blue indicates RADAR-WMH and orange indicates WHITE-Net. Asterisks denote FDR-corrected significant effects (pFDR < 0.05), and black triangles denote significant interaction effects. WMH volume corresponds to log-transformed lesion volume. SBP and cognitive analyses shown here were not adjusted for age; corresponding age-adjusted analyses are provided in **Supplementary Figure 1**.

We next investigated whether WMH-derived MRI metrics obtained from each segmentation method captured biologically meaningful variation related to aging, vascular burden, and cognition, and whether one method provided greater sensitivity to these clinically relevant variables. To address this, we assessed both the strength of associations between MRI metrics and clinical measures (β coefficients), and the additional predictive value provided by each metric beyond demographic covariates (ΔR²).

Age was strongly associated with multiple WMH-derived MRI metrics for both methods (Figure 6B). Across approaches, older age was associated with lower R2* values and higher ISOVF and MD values. Several metrics showed significant method-by-age interaction effects, with RADAR-derived metrics demonstrating stronger age-related decreases in MTsat, R1 and g-ratio, whereas WHITE-Net-derived WMH volume showed a stronger positive association with age. Adding WMH-derived MRI metrics to demographic covariates improved age prediction, with the largest ΔR² values observed for WMH volume and ISOVF. For most metrics, including MTsat, R1, R2*, FA, OD, g-ratio, ISOVF, and MD, RADAR-derived measures provided greater additional predictive value than corresponding WHITE-Net-derived measures, whereas WHITE-Net-derived WMH volume showed higher predictive value.

Associations with SBP were overall weaker but followed a similar pattern (Figure 6C). Higher SBP was associated with larger WMH volume and lower OD values across both methods. RADAR-derived metrics additionally showed an association between higher SBP and lower MTsat, R1 and higher ISOVF values. The additional predictive value of WMH-derived MRI metrics for SBP was modest overall, and although some differences between methods reached statistical significance, effect sizes were small. For cognitive performance, poorer TMT-A performance was associated with a microstructural pattern that closely mirrored the pattern observed with increasing age (Figure 6D, left). A similar, albeit weaker, pattern was observed for TMT-B performance (Figure 6D, right). In both cases, associations were consistently stronger for RADAR-derived than for WHITE-Net-derived microstructural measures, suggesting that RADAR-WMH captures tissue alterations that are more relevant to cognitive function.

## Discussion

Current WMH segmentation methods implicitly assume that FLAIR-visible lesions provide a comprehensive representation of clinically relevant white matter injury (Cathala et al. 2025; Gesierich et al. 2025; Gibson et al. 2026; Park et al. 2021; Puonti et al. 2016). Our findings challenge this assumption - a brain microstructure-based segmentation approach identified tissue abnormalities that overlapped with, but were not fully captured by, conventional FLAIR-defined WMH. Despite being invisible to standard lesion definitions, the regions identified exclusively by RADAR-WMH exhibit microstructural signatures consistent with myelin loss and axonal degeneration, remained stable over time, and were associated with age, vascular risk, and cognition, indicating that they represent a meaningful component of white matter injury rather than segmentation noise.

The best performing RADAR-WMH model integrated voxel-wise quantitative MRI metrics including local neighbouring voxel and textural information. This likely reflects the biological organisation of WM lesion progression, which occurs predominantly through expansion of existing lesions rather than emergence of new isolated foci (Maillard et al. 2012; Fazekas et al. 1993). Moreover, tissue surrounding visible WMH exhibits measurable microstructural pathology and increased risk of future lesion progression - a phenomenon commonly termed the WMH “penumbra” (Maillard et al. 2014-6; Maniega et al. 2015; Voorter et al. 01/2025; Bussy et al. 2025). Lesion voxels are therefore expected to be embedded within a broader region of tissue injury, making neighbourhood information particularly informative for segmentation.

One advantage of LGBM-based segmentation is its inherent interpretability - feature importance metrics and SHAP values allow transparent identification of the features driving model predictions. Diffusion-derived metrics showed the strongest contribution to lesion classification, with MD and ISOVF the most relevant, followed by OD and ICVF. This is consistent with previous imaging studies showing differences in these metrics in WMH relative to NAWM (Maniega et al. 2015; Iordanishvili et al. 2019; Jochems et al. 2024; Bastin et al. 2009; O’Sullivan et al. 2004; Bussy et al. 2025).

The strong contribution of MD and ISOVF likely reflects their sensitivity to extracellular free water accumulation, one of the earliest and most prominent microstructural changes associated with cerebral small vessel disease (Duering et al. 2018; Voorter et al. 01/2025). Increased free water tissue content may arise through several mechanisms, including blood– brain barrier dysfunction, periventricular fluid transudation, and tissue rarefaction secondary to axonal and oligodendroglial loss (Gouw et al. 2011; Beach et al. 2023; Masumura et al. 2001; Shim et al. 2015; Bridges et al. 2014)

Relaxometry-derived metrics contributed less strongly to classification despite their known sensitivity to myelin and iron content (Stüber et al. 2014). One possible explanation is that demyelination extends beyond the boundaries of FLAIR-visible lesions, making it less spatially specific than free water accumulation (Bronge et al. 2002; Fernando et al. 2004; Roseborough et al. 2020; Grafton et al. 1991). Relaxometry measures also reflect both myelin loss and iron accumulation, which may exert opposing effects on MRI contrast and thereby reduce lesion conspicuity (Gebril et al. 2011; Adeniyi et al. 2023). Together, these findings suggest that diffusion-derived metrics provide the strongest signal for identifying tissue corresponding to conventional WMH, while relaxometry-derived measures contribute complementary information on tissue composition.

Examination of segmentation errors and uncertainty provided further insight into the biological characteristics of tissue near lesion boundaries. Voxels missed by RADAR-WMH showed less severe microstructural abnormalities than voxels included in the manually segmented WMH ground truth and were predominantly located within smaller lesions. Similarly, uncertainty was highest at lesion borders and in small lesions, consistent with a continuum between NAWM and the lesion core (Maillard et al. 2014-6; Maniega et al. 2015; Bussy et al. 2025; Silbert et al. 2024; Voorter et al. 01/2025). These findings suggest that uncertainty reflects biologically intermediate tissue rather than model failure. Moreover, participant-level uncertainty correlated with segmentation performance, supporting its potential utility as a surrogate measure of segmentation quality when no ground truth is available (Philps et al. 2025).

A central objective of this study was to determine whether microstructure-based lesion segmentation identifies tissue abnormalities beyond those captured by conventional FLAIR imaging. Longitudinal comparisons between RADAR-WMH and WHITE-Net indicate that the two approaches are sensitive to partially distinct aspects of white matter pathology. RADAR-WMH-only voxels showed microstructural abnormalities highly similar to those detected by both methods, whereas WHITE-Net-only voxels showed less pathology, with a profile closer to NAWM. This pattern is consistent with the microstructure-driven nature of RADAR-WMH segmentation, and with pathological (Fazekas et al. 1993; Udaka and Sawada 2002; Wardlaw et al. 2015; Gouw et al. 2011) and in-vivo (Bussy et al. 2025; Parent et al. 2023) evidence that more severe tissue damage corresponds to more advanced lesion stages.

Notably, these differences could not be explained solely by FLAIR intensity alone - RADAR-WMH-only voxels showed higher FLAIR signal intensity than WHITE-Net-only voxels despite markedly greater microstructural pathology. This is consistent with the recognised microstructural heterogeneity of FLAIR-defined lesions (Gouw et al. 2011; Wardlaw et al. 2015) and suggest that lesion appearance on FLAIR is only partially related to the severity of underlying tissue damage. Areas with substantial microstructural impairment may remain undetected by FLAIR-based segmentation, while FLAIR-visible abnormalities can reflect relatively mild tissue pathology.

Longitudinal analyses further supported the biological relevance of RADAR-WMH-only detected lesions. These voxels showed greater temporal stability than WHITE-Net-only voxels, with nearly two-thirds remaining RADAR-specific at follow-up compared with less than half of WHITE-NET-only voxels remaining WHITE-Net-specific. Within RADAR-WMH-only voxels, the degree of microstructural abnormality appeared to predict persistence - voxels that remained RADAR-WMH-only showed the most severe tissue pathology, whereas those that disappeared showed comparatively milder abnormalities. This is consistent with previous reports that regressing WMH retain relatively preserved microstructural properties compared with persistent lesions (Jochems et al. 2024; Maillard et al. 2012). Notably, even RADAR-WMH-only voxels that disappeared at follow-up remained more microstructurally abnormal than persistent WHITE-Net-only voxels. Persistent RADAR-WMH-only voxels also showed higher FLAIR intensity than persistent WHITE-Net-only voxels, indicating that these differences cannot be explained by conventional lesion contrast alone.

Together, these findings suggest that FLAIR-based and microstructure-based segmentation identify partially distinct tissue compartments rather than merely differing lesion boundaries. The close resemblance between RADAR-WMH-only and jointly detected voxels, combined with their longitudinal stability, argues against a simple false-positive interpretation and indicates that RADAR-WMH captures a biologically meaningful component of white matter injury - characterized by increased extracellular water, myelin loss and axonal degeneration - that is incompletely represented by conventional FLAIR imaging.

The analyses above indicate that FLAIR-based and microstructure-based segmentations are sensitive to different aspects of white matter pathology. FLAIR-based segmentation captures areas of abnormal signal intensity with heterogeneous, sometimes mild microstructural impairment, while missing areas with clear microstructural damage. The key question is therefore what added clinical value a microstructure-based approach offers over FLAIR-based segmentation. Our data suggest a nuanced answer. FLAIR-based lesion volume was slightly more predictive of age and cognitive outcomes such as processing speed and executive function, however, the largest differences favoured RADAR-WMH, with MTsat, R1, ISOVF and MD measured within RADAR-WMH-segmented lesions showing greater predictive power for age and cognition than the same metrics measured within FLAIR-defined lesions. This likely reflects RADAR-WMH’s sensitivity to diffuse increases in extracellular free water and myelin-related signal changes extending beyond discrete FLAIR visible lesions and is consistent with evidence that both WMH and NAWM microstructural differences carry predictive value beyond lesion burden alone (Tuladhar et al. 2015; van Norden et al. 2012; Vernooij et al. 2009).

Our study has several limitations. First, manually segmented FLAIR-defined WMH served as the training and evaluation ground truth. While these remain the reference standard in the field, they are inherently constrained by lesion detectability on a single MRI contrast and may not fully capture the spectrum of underlying white matter pathology. Consequently, microstructurally abnormal tissue identified by RADAR-WMH but absent from the manual reference is penalised as a false positive under conventional performance metrics, potentially underestimating the value of microstructure-based segmentation. Second, discrepancies between RADAR-WMH and WHITE-Net may partly reflect limitations of WHITE-Net rather than to FLAIR itself, although WHITE-Net performs comparably to other state-of-the-art FLAIR-based algorithms (Cathala et al. 2025), suggesting that the observed differences are unlikely to be specific to a single method. More broadly, segmentation based on conventional MRI protocols is inherently limited by reliance on unimodal information, a constraint shared by human raters (Kuijf et al. 11/2019), which multi-contrast approaches such as RADAR-WMH may help overcome. Finally, the longitudinal sample was relatively small and included only a single follow-up timepoint, limiting conclusions regarding long-term lesion evolution and whether RADAR-only lesions eventually become FLAIR-visible or represent a distinct stage of white matter injury.

Our findings suggest that microstructure-based segmentation captures a clinically meaningful burden of white matter injury that remains invisible to conventional FLAIR-based approaches. RADAR-WMH may therefore offer a more sensitive and biologically grounded tool for tracking white matter disease and evaluating intervention effects in cerebral small vessel disease.

## Data availability statement

Data from BrainLaus dataset is not publicly available but can be requested upon reasonable and formal request (https://www.colaus-psycolaus.ch/professionals/how-to-collaborate). The code can be found on <u>GitHub</u> and the tool is publicly available on <u>Google Drive</u>.

## Acknowledgments

We wish to thank all BrainLaus participants for their participation in the study. ChatGPT (OpenAI) was used solely for language editing. All scientific content and references were reviewed and verified by the authors, who take full responsibility for the manuscript.

## Funding

B.D. is supported by the Swiss National Science Foundation (project grant no. 213595, 32003B_135679, 32003B_159780, 324730_192755 and CRSK-3_190185), InnoSuisse Flagship Swiss brAInHealth project, ERA_NET NEURON JTC2020: iSEE and JTC2023-ELSA: BrainTree projects. F.K. is funded by the PHASE IV AI/Horizon Europe grant, grant agreement ID: 1010953844:01 and Horizon 2020 (871643-MORPHEMIC). A.L. is supported by the Swiss National Science Foundation (Grant Nos. 320030_184784, CR00I5-235940). The Laboratory for Research in Neuroimaging (LREN) is very grateful to the Roger De Spoelberch and Partridge Foundations for their generous financial support. The CoLaus|PsyCoLaus study was supported by unrestricted research grants from GlaxoSmithKline, the Faculty of Biology and Medicine of Lausanne, the Swiss National Science Foundation (grants 3200B0–105993, 3200B0-118308, 33CSCO-122661, 33CS30-139468, 33CS30-148401, 33CS30_177535, 324730_204523 and 320030_220190) and the Swiss Personalized Health Network (grant 2018DRI01).

## Competing interests

The authors report no competing interests.

## Notes

### Competing Interest Statement

The authors have declared no competing interest.

### Author Declarations

Ethical approval was granted by the Ethics Commission of Canton de Vaud, and participants gave their informed consent prior to participation.

## References

Adeniyi, P. A., X. Gong, E. MacGregor, et al. 2023. “Ferroptosis of Microglia in Aging Human White Matter Injury.” Annals of Neurology 94 (6): 1048–1066. 37605362.

Akiba, Takuya, Shotaro Sano, Toshihiko Yanase, Takeru Ohta, and Masanori Koyama. 2019. “Optuna: A next-Generation Hyperparameter Optimization Framework.” Proceedings of the 25th ACM SIGKDD International Conference on Knowledge Discovery & Data Mining, July 25, 2623–2631.

Andersson, Jesper L. R., and Stamatios N. Sotiropoulos. 2016. “An Integrated Approach to Correction for off-Resonance Effects and Subject Movement in Diffusion MR Imaging.” NeuroImage 125 (January): 1063–1078.

Ashburner, John, and Karl J. Friston. 2005. “Unified Segmentation.” NeuroImage 26 (3): 839–851.

Bastin, Mark E., Jonathan D. Clayden, Alison Pattie, Iona F. Gerrish, Joanna M. Wardlaw, and Ian J. Deary. 2009. “Diffusion Tensor and Magnetization Transfer MRI Measurements of Periventricular White Matter Hyperintensities in Old Age.” Neurobiology of Aging 30 (1): 125–136.

Beach, Thomas G., Lucia I. Sue, Sarah Scott, et al. 2023. “Cerebral White Matter Rarefaction Has Both Neurodegenerative and Vascular Causes and May Primarily Be a Distal Axonopathy.” Journal of Neuropathology and Experimental Neurology 82 (6): 457–466.

Bridges, L. R., J. Andoh, A. J. Lawrence, et al. 2014. “Blood-Brain Barrier Dysfunction and Cerebral Small Vessel Disease (arteriolosclerosis) in Brains of Older People.” Journal of Neuropathology and Experimental Neurology 73 (11): 1026–1033. 25289893.

Bronge, Lena, Nenad Bogdanovic, and Lars-Olof Wahlund. 2002. “Postmortem MRI and Histopathology of White Matter Changes in Alzheimer Brains: A Quantitative, Comparative Study.” Dementia and Geriatric Cognitive Disorders 13 (4): 205–212.

Bussy, Aurélie, Camille Cathala, Fábio Carneiro, Ferath Kherif, Antoine Lutti, and Bogdan Draganski. 2025. “White Matter Microstructure Fingerprint of Cerebral Small Vessel Disease.” In bioRxiv. 10.1101/2025.10.01.679737.

Cathala, Camille, Ferath Kherif, Jean-Philippe Thiran, Aurélie Bussy, and Bogdan Draganski. 2025. “WHITE-Net : White Matter HyperIntensities Tissue Extraction Using Deep Learning Network.” In medRxiv. January 9. 10.1101/2025.01.09.25320242.

Charidimou, Andreas, Gregoire Boulouis, Matthew P. Frosch, et al. 2022. “The Boston Criteria Version 2.0 for Cerebral Amyloid Angiopathy: A Multicentre, Retrospective, MRI–neuropathology Diagnostic Accuracy Study.” Lancet Neurology 21 (8): 714–725.

Daducci, Alessandro, Erick J. Canales-Rodríguez, Hui Zhang, Tim B. Dyrby, Daniel C. Alexander, and Jean-Philippe Thiran. 2015. “Accelerated Microstructure Imaging via Convex Optimization (AMICO) from Diffusion MRI Data.” NeuroImage 105 (January): 32–44.

Draganski, B., J. Ashburner, C. Hutton, et al. 2011. “Regional Specificity of MRI Contrast Parameter Changes in Normal Ageing Revealed by Voxel-Based Quantification (VBQ).” NeuroImage 55 (4): 1423–1434.

Duering, Marco, Geert Jan Biessels, Amy Brodtmann, et al. 2023. “Neuroimaging Standards for Research into Small Vessel Disease-Advances since 2013.” The Lancet. Neurology 22 (7): 602–618.

Duering, Marco, Sofia Finsterwalder, Ebru Baykara, et al. 2018. “Free Water Determines Diffusion Alterations and Clinical Status in Cerebral Small Vessel Disease.” Alzheimer’s & Dementia : The Journal of the Alzheimer’s Association 14 (6): 764–774.

Fazekas, F., R. Kleinert, H. Offenbacher, et al. 1993. “Pathologic Correlates of Incidental MRI White Matter Signal Hyperintensities.” Neurology 43 (9): 1683–1689.

Fernando, M. S., J. T. O’Brien, R. H. Perry, et al. 2004. “Comparison of the Pathology of Cerebral White Matter with Post-Mortem Magnetic Resonance Imaging (MRI) in the Elderly Brain.” Neuropathology and Applied Neurobiology 30 (4): 385–395. 15305984.

Firmann, Mathieu, Vladimir Mayor, Pedro Marques Vidal, et al. 2008. “The CoLaus Study: A Population-Based Study to Investigate the Epidemiology and Genetic Determinants of Cardiovascular Risk Factors and Metabolic Syndrome.” BMC Cardiovascular Disorders 8 (March): 6.

Gebril, O. H., J. E. Simpson, J. Kirby, C. Brayne, and P. G. Ince. 2011. “Brain Iron Dysregulation and the Risk of Ageing White Matter Lesions.” Neuromolecular Medicine 13(4): 289–299. 51650443.

Gesierich, Benno, Lukas Pirpamer, Dominik S. Meier, et al. 2025. “Technical and Clinical Validation of a Novel Deep Learning-Based White Matter Hyperintensity Segmentation Tool.” Cerebral Circulation - Cognition and Behavior 9 (100393): 100393.

Gibson, Erin, Joel Ramirez, Lauren Abby Woods, et al. 2026. “SegcsvdPVS: A Convolutional Neural Network-Based Tool for Quantification of Enlarged Perivascular Spaces (PVS) on T1-Weighted Images.” Human Brain Mapping 47 (2): e70462.

Gouw, Alida A., Alexandra Seewann, Wiesje M. Van Der Flier, et al. 2011. “Heterogeneity of Small Vessel Disease: A Systematic Review of MRI and Histopathology Correlations.” Journal of Neurology, Neurosurgery, and Psychiatry 82 (2): 126–135.

Grafton, S. T., S. M. Sumi, G. K. Stimac, E. C. Alvord Jr, C. M. Shaw, and D. Nochlin. 1991. “Comparison of Postmortem Magnetic Resonance Imaging and Neuropathologic Findings in the Cerebral White Matter.” Archives of Neurology 48 (3): 293–298. 1705796.

Haddad, Seyyed M. H., Christopher J. M. Scott, Miracle Ozzoude, et al. 2022. “Comparison of Diffusion Tensor Imaging Metrics in Normal-Appearing White Matter to Cerebrovascular Lesions and Correlation with Cerebrovascular Disease Risk Factors and Severity.” International Journal of Biomedical Imaging 2022 (October): 5860364.

Haight, Thaddeus, R. Nick Bryan, Guray Erus, et al. 2018. “White Matter Microstructure, White Matter Lesions, and Hypertension: An Examination of Early Surrogate Markers of Vascular-Related Brain Change in Midlife.” NeuroImage. Clinical 18 (March): 753–761.

Hassija, Vikas, Vinay Chamola, Atmesh Mahapatra, et al. 2024. “Interpreting Black-Box Models: A Review on Explainable Artificial Intelligence.” Cognitive Computation 16 (1): 45–74.

Iordanishvili, Elene, Melissa Schall, Ricardo Loução, et al. 2019. “Quantitative MRI of Cerebral White Matter Hyperintensities: A New Approach towards Understanding the Underlying Pathology.” NeuroImage 202 (November): 116077.

James, Sarah-Naomi, Emily N. Manning, Mathew Storey, et al. 2023. “Neuroimaging, Clinical and Life Course Correlates of Normal-Appearing White Matter Integrity in 70-Year-Olds.” Brain Communications 5 (5): fcad225.

Jenkinson, Mark, Peter Bannister, Michael Brady, and Stephen Smith. 2002. “Improved Optimization for the Robust and Accurate Linear Registration and Motion Correction of Brain Images.” NeuroImage 17 (2): 825–841.

Jochems, Angela C. C., Susana Muñoz Maniega, Una Clancy, et al. 2024. “Magnetic Resonance Imaging Tissue Signatures Associated With White Matter Changes Due to Sporadic Cerebral Small Vessel Disease Indicate That White Matter Hyperintensities Can Regress.” Journal of the American Heart Association 13 (3): e032259.

Kort, Floor A. S. de, Elisabeth J. Vinke, Ewoud J. van der Lelij, et al. 2025. “Cerebral White Matter Hyperintensity Volumes: Normative Age- and Sex-Specific Values from 15 Population-Based Cohorts Comprising 14,876 Individuals.” Neurobiology of Aging 146 (February): 38–47.

Kuijf, Hugo J., Adria Casamitjana, D. Louis Collins, et al. 11/2019. “Standardized Assessment of Automatic Segmentation of White Matter Hyperintensities and Results of the WMH Segmentation Challenge.” IEEE Transactions on Medical Imaging 38 (11): 2556–2568.

Leijsen, Esther M. C. van, Mayra I. Bergkamp, Ingeborg W. M. van Uden, et al. 2018. “Progression of White Matter Hyperintensities Preceded by Heterogeneous Decline of Microstructural Integrity.” Stroke 49 (6): 1386–1393.

Lundberg, S. M., and S. -. I. Lee. 2017. A Unified Approach to Interpreting Model Predictions”. Curran Associates, Inc.

Lutti, Antoine, Frederic Dick, Martin I. Sereno, and Nikolaus Weiskopf. 2014. “Using High-Resolution Quantitative Mapping of R1 as an Index of Cortical Myelination.” NeuroImage 93: 176–188.

Lutti, Antoine, Chloe Hutton, Jürgen Finsterbusch, Gunther Helms, and Nikolaus Weiskopf. 2010. “Optimization and Validation of Methods for Mapping of the Radiofrequency Transmit Field at 3T.” Magnetic Resonance in Medicine: Official Journal of the Society of Magnetic Resonance in Medicine / Society of Magnetic Resonance in Medicine 64 (1): 229–238.

Lutti, Antoine, Joerg Stadler, Oliver Josephs, et al. 2012. “Robust and Fast Whole Brain Mapping of the RF Transmit Field B1 at 7T.” PloS One 7 (3): e32379.

Maier-Hein, Lena, Matthias Eisenmann, Annika Reinke, et al. 2018. “Why Rankings of Biomedical Image Analysis Competitions Should Be Interpreted with Care.” Nature Communications 9 (1): 5217.

Maillard, Pauline, Owen Carmichael, Evan Fletcher, Bruce Reed, Dan Mungas, and Charles DeCarli. 2012. “Coevolution of White Matter Hyperintensities and Cognition in the Elderly.” Neurology 79 (5): 442–448.

Maillard, Pauline, Evan Fletcher, Sam N. Lockhart, et al. 2014-6. “White Matter Hyperintensities and Their Penumbra Lie Along a Continuum of Injury In The Aging Brain.” Stroke; a Journal of Cerebral Circulation 45 (6): 1721–1726.

Maillard, P., O. Carmichael, D. Harvey, et al. 2013. “FLAIR and Diffusion MRI Signals Are Independent Predictors of White Matter Hyperintensities.” AJNR. American Journal of Neuroradiology 34 (1): 54–61.

Maniega, Susana Muñoz, Maria C. Valdés Hernández, Jonathan D. Clayden, et al. 2015. “White Matter Hyperintensities and Normal-Appearing White Matter Integrity in the Aging Brain.” Neurobiology of Aging 36 (2): 909–918.

Masumura, M., R. Hata, H. Akatsu, et al. 2001. “Increasing in Situ Nick End Labeling of Oligodendrocytes in White Matter of Patients with Binswanger’s Disease.” Journal of Stroke and Cerebrovascular Diseases: The Official Journal of National Stroke Association 10(2): 55–62. 32489095.

Mitchell, Rory, Eibe Frank, and Geoffrey Holmes. 2022. “GPUTreeShap: Massively Parallel Exact Calculation of SHAP Scores for Tree Ensembles.” PeerJ. Computer Science 8 (e880): e880.

Muñoz Maniega, Susana, Francesca M. Chappell, Maria C. Valdés Hernández, et al. 2017. “Integrity of Normal-Appearing White Matter: Influence of Age, Visible Lesion Burden and Hypertension in Patients with Small-Vessel Disease.” Journal of Cerebral Blood Flow and Metabolism : Official Journal of the International Society of Cerebral Blood Flow and Metabolism 37 (2): 644–656.

Norden, Anouk G. W. van, Karlijn F. de Laat, Ewoud J. van Dijk, et al. 2012. “Diffusion Tensor Imaging and Cognition in Cerebral Small Vessel Disease: The RUN DMC Study.” Biochimica et Biophysica Acta 1822 (3): 401–407.

Ojala, Timo, Matti Pietikäinen, and David Harwood. 1996. “A Comparative Study of Texture Measures with Classification Based on Featured Distributions.” Pattern Recognition 29 (1): 51–59.

O’Sullivan, M., R. G. Morris, B. Huckstep, D. K. Jones, S. C. R. Williams, and H. S. Markus. 2004. “Diffusion Tensor MRI Correlates with Executive Dysfunction in Patients with Ischaemic Leukoaraiosis.” Journal of Neurology, Neurosurgery, and Psychiatry 75 (3): 441–447.

Parent, Olivier, Aurélie Bussy, Gabriel Allan Devenyi, et al. 2023. “Assessment of White Matter Hyperintensity Severity Using Multimodal Magnetic Resonance Imaging.” Brain Communications 5 (6): 1–18.

Park, Gilsoon, Jinwoo Hong, Ben A. Duffy, Jong-Min Lee, and Hosung Kim. 2021. “White Matter Hyperintensities Segmentation Using the Ensemble U-Net with Multi-Scale Highlighting Foregrounds.” NeuroImage 237 (118140): 118140.

Philps, Ben, Maria Del C. Valdés Hernández, Chen Qin, et al. 2025. “Uncertainty Quantification for White Matter Hyperintensity Segmentation Detects Silent Failures and Improves Automated Fazekas Quantification.” Medical Image Analysis 105 (October): 103697.

Puonti, Oula, Juan Eugenio Iglesias, and Koen Van Leemput. 2016. “Fast and Sequence-Adaptive Whole-Brain Segmentation Using Parametric Bayesian Modeling.” NeuroImage 143 (December): 235–249.

Rainio, Oona, and Riku Klén. 2026. “Modified Dice Coefficients for Evaluation of Tumor Segmentation from PET Images: A Proof-of-Concept Study.” Journal of Imaging Informatics in Medicine 39 (1): 785–793.

Roseborough, Austyn D., Kristopher D. Langdon, Robert Hammond, et al. 2020. “Post-Mortem 7 Tesla MRI Detection of White Matter Hyperintensities: A Multidisciplinary Voxel-Wise Comparison of Imaging and Histological Correlates.” NeuroImage. Clinical 27 (January): 102340.

Shim, Yong S., Dong-Won Yang, Catherine M. Roe, et al. 2015. “Pathological Correlates of White Matter Hyperintensities on MRI.” Dementia and Geriatric Cognitive Disorders 39 (0): 92–104.

Silbert, Lisa C., Natalie E. Roese, Victoria Krajbich, et al. 2024. “White Matter Hyperintensities and the Surrounding Normal Appearing White Matter Are Associated with Water Channel Disruption in the Oldest Old.” Alzheimer’s & Dementia: The Journal of the Alzheimer’s Association 20 (6): 3839–3851.

Simpson, J. E., O. Hosny, S. B. Wharton, et al. 2009. “Microarray RNA Expression Analysis of Cerebral White Matter Lesions Reveals Changes in Multiple Functional Pathways.” Stroke; a Journal of Cerebral Circulation 40 (2): 369–375. 19109541.

Simpson, J. E., P. G. Ince, C. E. Higham, et al. 2007. “Microglial Activation in White Matter Lesions and Nonlesional White Matter of Ageing Brains.” Neuropathology and Applied Neurobiology 33 (6): 670–683. 17990995.

Slater, David A., Lester Melie-Garcia, Martin Preisig, Ferath Kherif, Antoine Lutti, and Bogdan Draganski. 2019. “Evolution of White Matter Tract Microstructure across the Life Span.” Human Brain Mapping 40 (7): 2252–2268.

Smith, Eric E., Gustavo Saposnik, Geert Jan Biessels, et al. 2017. “Prevention of Stroke in Patients With Silent Cerebrovascular Disease: A Scientific Statement for Healthcare Professionals From the American Heart Association/American Stroke Association.” Stroke 48 (2): e44–e71.

Solé-Guardia, Gemma, Matthijs Luijten, Esther Janssen, et al. 2025. “Deep Learning-Based Segmentation in MRI-(immuno)histological Examination of Myelin and Axonal Damage in Normal-Appearing White Matter and White Matter Hyperintensities.” Brain Pathology (Zurich, Switzerland) 35 (2): e13301.

Stikov, Nikola, Jennifer S. W. Campbell, Thomas Stroh, et al. 2015. “In Vivo Histology of the Myelin G-Ratio with Magnetic Resonance Imaging.” NeuroImage 118 (September): 397–405.

Stüber, Carsten, Markus Morawski, Andreas Schäfer, et al. 2014. “Myelin and Iron Concentration in the Human Brain: A Quantitative Study of MRI Contrast.” NeuroImage 93: 95–106.

Tofts, Paul. 2003. Quantitative MRI of the Brain: Measuring Changes Caused by Disease. John Wiley & Sons.

Tournier, J-Donald, Robert Smith, David Raffelt, et al. 2019. “MRtrix3: A Fast, Flexible and Open Software Framework for Medical Image Processing and Visualisation.” NeuroImage 202 (November): 116137.

Trofimova, Olga, Leyla Loued-Khenissi, Giulia DiDomenicantonio, et al. 2021. “Brain Tissue Properties Link Cardio-Vascular Risk Factors, Mood and Cognitive Performance in the CoLaus|PsyCoLaus Epidemiological Cohort.” Neurobiology of Aging 102 (June): 50–63.

Tuladhar, Anil M., Anouk G. W. van Norden, Karlijn F. de Laat, et al. 2015. “White Matter Integrity in Small Vessel Disease Is Related to Cognition.” NeuroImage. Clinical 7 (February): 518–524.

Udaka, F., and H. Sawada. 2002. “White Matter Lesions and Dementia MRI pathological Correlation.” Annals of the New York …, ahead of print. 10.1111/j.1749-6632.2002.tb04845.x.

Vernooij, Meike W., M. Arfan Ikram, Henri A. Vrooman, et al. 2009. “White Matter Microstructural Integrity and Cognitive Function in a General Elderly Population.” Archives of General Psychiatry 66 (5): 545–553.

Voorter, Paulien H. M., Michael S. Stringer, Maud Van Dinther, et al. 01/2025. “Heterogeneity and Penumbra of White Matter Hyperintensities in Small Vessel Diseases Determined by Quantitative MRI.” Stroke; a Journal of Cerebral Circulation 56 (1): 128–137.

Wardlaw, Joanna M., Maria C. Valdés Hernández, and Susana Muñoz-Maniega. 2015. “What Are White Matter Hyperintensities Made Of?” Journal of the American Heart Association 4 (6): e001140.

Weiskopf, Nikolaus, John Suckling, Guy Williams, et al. 2013. “Quantitative Multi-Parameter Mapping of R1, PD(*), MT, and R2(*) at 3T: A Multi-Center Validation.” Frontiers in Neuroscience 7 (June): 95.

Zhang, Hui, Torben Schneider, Claudia A. Wheeler-Kingshott, and Daniel C. Alexander. 2012. “NODDI: Practical in Vivo Neurite Orientation Dispersion and Density Imaging of the Human Brain.” NeuroImage 61 (4): 1000–1016.

