## Supplementary material for "RADAR-WMH: Relaxometry And Diffusion Analysis beyond Radiologically defined WMH"

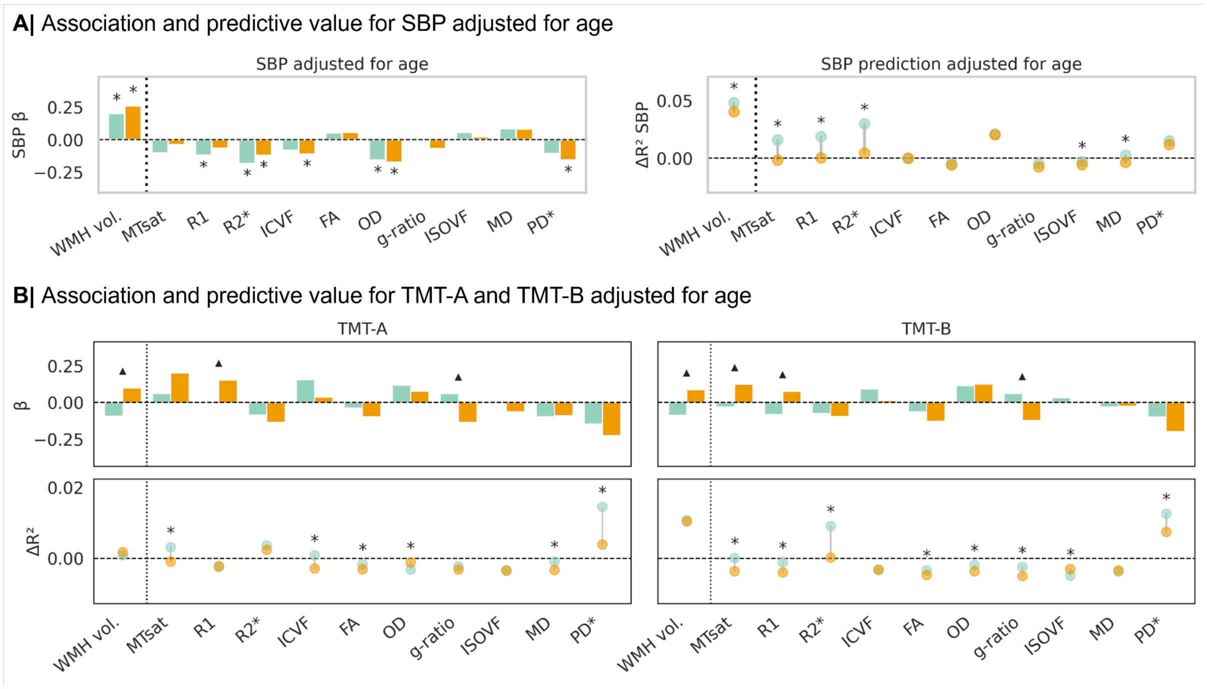


**Supplementary figure 1:** Association and predictive value analyses for systolic blood pressure (SBP; **A**), and cognitive performance measured using Trail Making Test A and B (TMT-A and TMT-B; **B**), analyses adjusted for age, sex, and education. Upper panels show standardized regression coefficients (β) from paired linear models relating MRI metrics to each clinical variable. Lower panels show the additional cross-validated predictive value ($\Delta R^2$) obtained when adding each MRI metric to a baseline model including age, sex and education, reflecting variance explained beyond covariates alone. $\Delta R^2$ values from RADAR- and WHITE-Net-derived metrics were directly compared using permutation testing. Blue indicates RADAR-WMH and orange indicates WHITE-Net. Asterisks denote FDR-corrected significant effects ($p_{FDR} < 0.05$), and black triangles denote significant interaction effects. WMH volume corresponds to log-transformed lesion volume.

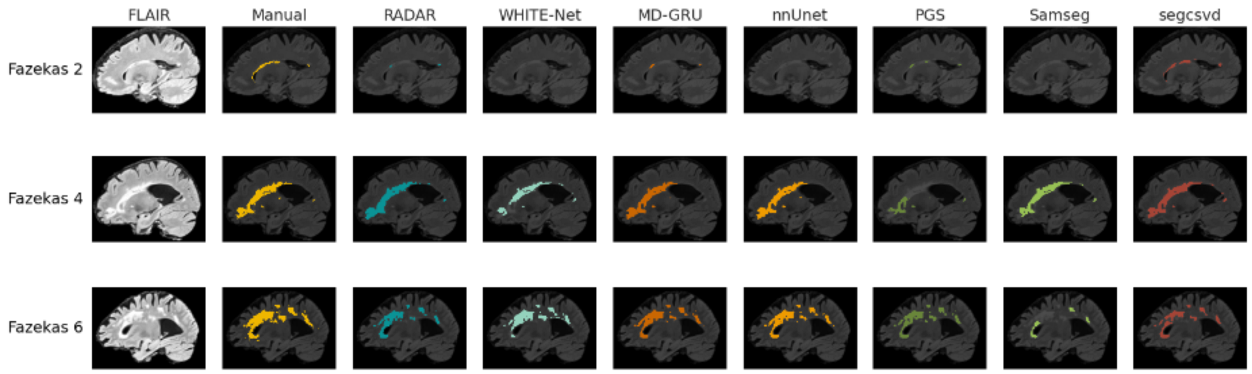


**Supplementary figure 2:** FLAIR images, manual segmentations, and binary prediction maps for all models in three example cases (Fazekas scores 2, 4, and 6), sagittal view.

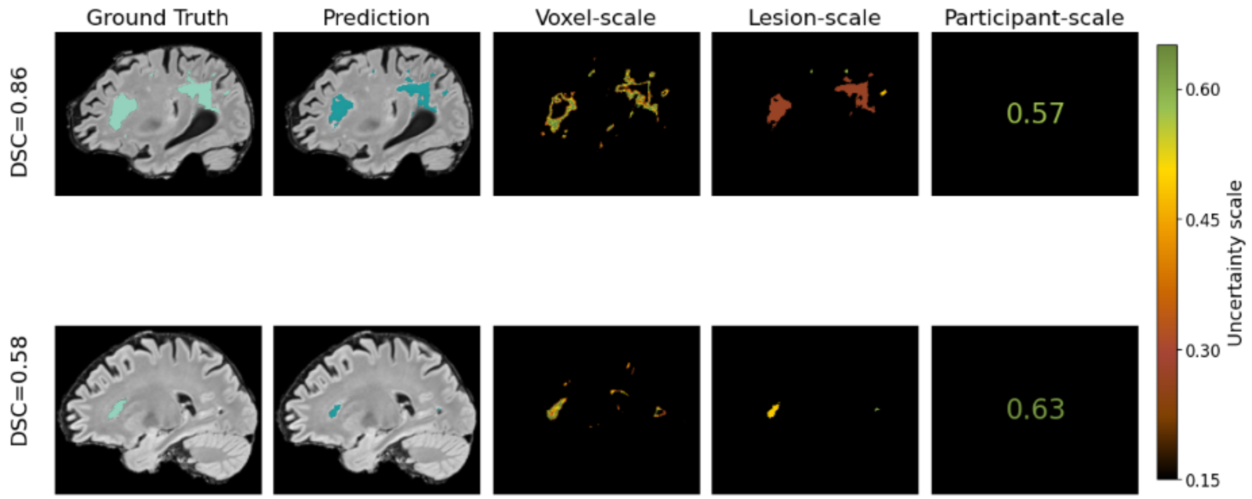


**Supplementary figure 3:** Sagittal views of uncertainty maps at multiple scales. The first column shows manual segmentations overlaid on FLAIR images, the second column shows predictions from the best-performing model, and the last three columns display uncertainty at the voxel-, lesion-, and participant-level, respectively

**Supplementary table 1**: Summary of parameter ranges for hyperparameter optimization of LGBM.

| **Parameters** | **Value range** |
|---|---|
| n_estimators | [100, 1000] |
| learning_rate | [0.01, 0.1] |

| | |
|---|---|
| num_leaves | [20, 100] |
| max_depth | [5, 30] |
| min_child_samples | [5, 100] |
| subsample | [0.6, 1.0] |
| colsample_bytree | [0.6, 1.0] |
| reg_alpha | [1e-8, 10.0] |
| reg_lambda | [1e-8, 10.0] |

**Supplementary table 2:** Performance comparison across models when Fazekas 0 is excluded. The mean value across participants with 95% bootstrap CI is presented. The best value is highlighted in **bold**.

| Model | DSC | Precision | SEN | VS | ASSD | RVD | HD95 | WDC |
|---|---|---|---|---|---|---|---|---|
| WHITE-Net | 0.54 [0.47, 0.61] | 0.58 [0.50, 0.65] | 0.56 [0.48, 0.62] | 0.78 [0.71, 0.83] | 8.02 [4.36, 12.58] | 0.61 [0.33, 1.03] | 20.04 [14.30, 26.74] | 0.58 [0.51, 0.65] |
| MD-GRU | 0.55 [0.48, 0.61] | 0.59 [0.51, 0.66] | 0.58 [0.53, 0.63] | 0.76 [0.69, 0.83] | 7.79 [4.11, 12.27] | 4.32 [0.56, 10.14] | 17.21 [11.48, 23.80] | 0.62 [0.55, 0.68] |
| nnUnet | **0.58 [0.52, 0.63]** | **0.63 [0.56, 0.69]** | **0.58 [0.53, 0.62]** | **0.81 [0.75, 0.86]** | **4.77 [1.92, 8.75]** | **0.57 [0.27, 1.06]** | **12.56 [8.07, 18.18]** | **0.66 [0.61, 0.71]** |
| segcsvd | 0.52 [0.45, 0.59] | 0.54 [0.46, 0.62] | **0.58 [0.52, 0.64]** | 0.71 [0.64, 0.79] | 7.03 [4.14, 10.50] | 2.31 [1.00, 3.99] | 18.06 [12.46, 24.57] | 0.57 [0.50, 0.64] |
| PGS | 0.51 [0.44, 0.56] | 0.54 [0.46, 0.61] | 0.56 [0.52, 0.60] | 0.72 [0.64, 0.78] | 6.61 [3.80, 10.18] | 1.72 [0.82, 2.85] | 17.92 [12.47, 24.39] | 0.59 [0.52, 0.65] |
| Shiva | 0.48 [0.42, 0.54] | 0.62 [0.54, 0.70] | 0.48 [0.42, 0.54] | 0.66 [0.60, 0.72] | 7.26 [3.76, 11.74] | 2.03 [0.57, 4.55] | 17.86 [12.52, 24.21] | 0.59 [0.53, 0.65] |
| Samseg | 0.46 [0.38, 0.53] | 0.53 [0.44, 0.61] | 0.43 [0.36, 0.50] | 0.66 [0.57, 0.75] | 5.68 [3.60, 8.36] | 1.13 [0.44, 2.31] | 18.12 [13.90, 22.66] | 0.48 [0.40, 0.55] |
| LGBM | 0.54 [0.47, 0.60] | 0.59 [0.50, 0.66] | 0.54 [0.48, 0.59] | 0.76 [0.69, 0.83] | 7.00 [4.10, 10.55] | 2.01 [0.56, 4.00] | 19.46 [13.66, 26.14] | 0.58 [0.51, 0.64] |

***Supplementary table 3:*** *Statistics for the Wilcoxon signed-rank test ($\boldsymbol{W}$)). Cliff's delta ($\boldsymbol{\delta}$) and FDR corrections ($W_{FDR}$ ) are also shown for each metric. Significant values after FDR are marked with a star (*)*

| **Comparison** | **MRI metric** | **Wilcoxon ($\boldsymbol{W}$)** | **Cliff's delta ($\boldsymbol{\delta}$)** | **Wilcoxon FDR $\boldsymbol{W_{FDR}}$** |
|---|---|---|---|---|
| Detected/Missed | MTsat | 1.45e-18 | -0.65 | 2.42e-18* |
| Detected/Missed | R1 | 2.10e-19 | -0.68 | 5.24e-19* |
| Detected/Missed | R2* | 8.75e-20 | -0.82 | 3.00e-19* |
| Detected/Missed | ICVF | 8.75e-20 | -0.76 | 3.00e-19* |
| Detected/Missed | FA | 2.98e-19 | -0.57 | 5.97e-19* |
| Detected/Missed | OD | 1.66e-13 | -0.44 | 2.37e-13* |
| Detected/Missed | g-ratio | 1.05e-02 | -0.12 | 1.05e-02* |
| Detected/Missed | ISOVF | 1.47e-03 | -0.16 | 1.63e-03* |
| Detected/Missed | MD | 9.23e-11 | 0.29 | 1.15e-10* |
| Detected/Missed | PD* | 8.99e-20 | 0.80 | 3.00e-19* |
| Missed/NAWM | MTsat | 1.55e-19 | -0.92 | 2.59e-19* |
| Missed/NAWM | R1 | 2.47e-19 | -0.84 | 3.52e-19* |
| Missed/NAWM | R2* | 1.00e-19 | -0.93 | 2.59e-19* |
| Missed/NAWM | ICVF | 8.99e-20 | -0.93 | 2.59e-19* |
| Missed/NAWM | FA | 8.23e-16 | -0.70 | 9.15e-16* |
| Missed/NAWM | OD | 1.21e-09 | -0.51 | 1.21e-09* |
| Missed/NAWM | g-ratio | 3.08e-16 | -0.63 | 3.85e-16* |
| Missed/NAWM | ISOVF | 1.22e-19 | 0.93 | 2.59e-19* |
| Missed/NAWM | MD | 9.50e-20 | 0.96 | 2.59e-19* |
| Missed/NAWM | PD* | 1.39e-19 | 0.98 | 2.59e-19* |
| Detected/NAWM | MTsat | 8.75e-20 | -0.99 | 1.32e-19* |
| Detected/NAWM | R1 | 8.75e-20 | -0.99 | 1.32e-19* |
| Detected/NAWM | R2* | 8.75e-20 | -1.00 | 1.32e-19* |
| Detected/NAWM | ICVF | 8.75e-19 | -0.99 | 1.32e-19* |
| Detected/NAWM | FA | 1.32e-19 | -0.94 | 1.65e-19* |
| Detected/NAWM | OD | 1.93e-17 | -0.81 | 2.14e-17* |
| Detected/NAWM | g-ratio | 5.30e-16 | -0.72 | 5.30e-16* |
| Detected/NAWM | ISOVF | 9.24e-20 | 0.96 | 1.32e-19* |
| Detected/NAWM | MD | 8.75e-19 | 0.99 | 1.32e-19* |
| Detected/NAWM | PD* | 8.75e-20 | 1.00 | 1.32e-19* |

### Supplementary methods

### MRI acquisition

#### *FLAIR*

FLAIR images were acquired with TR/TE/TI = 5000/389/1800 ms and 1-mm isotropic resolution. Acquisition time was reduced using GRAPPA acceleration (factor 2) in the phase-encoding direction and partial Fourier (7/8) in the slice direction.

#### *Relaxometry*

Relaxometry data were acquired using a custom 3D GRE pulse sequence (Weiskopf et al. 2013). Multi-echo images were obtained with T1-weighted (TR/α = 18.7 ms/20°), PD-weighted, and MT-weighted contrasts (TR/α = 23.7 ms/6°) with echo times ranging from 2.2 to 19.7 ms. Additional acquisition parameters included 1-mm isotropic resolution, matrix size 256 × 240 × 176, GRAPPA acceleration factor 2 in the phase-encoding direction, 6/8 partial Fourier in the partition direction, non-selective RF excitation, readout bandwidth 425 Hz/pixel, and RF spoiling phase increment 50°, resulting in a total acquisition time of approximately 19 minutes. B1 mapping was performed using the 3D EPI SE/STE method (Lutti et al. 2010, 2012) to correct for radio-frequency field inhomogeneities (Lutti et al. 2014).

#### *Diffusion*

Diffusion-weighted imaging (DWI) was acquired using a 2D echo-planar imaging (EPI) sequence with TR/TE = 7400/69 ms, GRAPPA acceleration factor 2, field of view 192 × 212 mm², voxel size 2 × 2 × 2 mm³, matrix size 96 × 106, and 70 axial slices. Diffusion encoding was performed along 118 directions (15 at b=650b=650b=650 s/mm², 30 at b=1000b=1000b=1000 s/mm², 60 at b=2000b=2000b=2000 s/mm²) with 13 interleaved b=0b=0b=0 images (Slater et al. 2019). B0 field maps were additionally acquired using a 2D double-echo FLASH sequence (slice thickness 2 mm, TR = 1020 ms, TE1/TE2 = 10/12.46 ms, flip angle 90°, bandwidth 260 Hz/pixel) and were used to correct geometric distortions in the EPI data.

### MRI preprocessing

#### Quantitative map estimation

Quantitative maps of magnetization transfer saturation (MTsat), the transverse relaxation rate (R2* = 1/T2*), the effective longitudinal relaxation rate (R1 = 1/T1), and the effective proton density (PD*) were computed from the raw MRI data using the VBQ toolbox (Draganski et al., 2011; Weiskopf et al., 2013). PD* maps were normalized assuming a mean value of 69% in white matter (Tofts 2003). This normalization removes inter-individual differences in PD* values across subjects while preserving intra-individual variability within the white matter, which can still reflect local tissue variations.

### Diffusion MRI processing

Diffusion-weighted imaging (DWI) data were preprocessed using MRtrix3 (Tournier et al., 2019), including denoising and removal of Gibbs ringing artifacts (Slater et al., 2019). Eddy current distortions and subject motion were corrected using the FSL 5.0 EDDY tool (Andersson and Sotiropoulos 2016). Susceptibility-induced distortions in the echo-planar imaging (EPI) data were corrected using the FieldMap toolbox of SPM12 with the acquired B0-field maps. Bias field correction was performed by estimating the bias field from the mean $b$=0 images and applying the correction to all diffusion-weighted volumes.

Diffusion tensor metrics were computed using the tensor model, yielding whole-brain maps of fractional anisotropy (FA) and mean diffusivity (MD). Multi-shell diffusion data were further analyzed using the neurite orientation dispersion and density imaging (NODDI) model (Zhang et al. 2012), implemented with the AMICO toolbox (Daducci et al. 2015), to derive maps of intracellular volume fraction (ICVF), isotropic volume fraction (ISOVF), and orientation dispersion index (OD).

MRI g-ratio maps, representing the ratio between the inner and outer diameters of the myelin sheath, were also computed (Slater et al., 2019; Stikov et al., 2015). Finally, all derived maps were aligned to the MTsat images using rigid-body registration implemented in SPM12.

### Feature descriptions

These different features and their mathematical descriptions are formally described in the following paragraphs.

#### 1. Local Features

The first group of features is known as Local Features. For a given kernel size $s$, a neighborhood is defined in a local square around the voxel of interest. From this neighborhood, two statistical descriptors are computed: the mean (Eq. 2) and the standard deviation (Eq. 3). Because the prediction task is performed voxel-wise, these two additional features provide contextual information about the surrounding environment to which the voxel belongs.

Let $N_r(i)$ denote the set of indices in a cubic neighborhood of radius $r$ around $i$.

$$N_r(i) = \{i + (u, v, w) \mid u, v, w \in [\![-r, r]\!]\} \qquad \text{(Eq. 1)}$$

Then we can define the mean ($X_i^{\mu}$) and standard deviation ($X_i^{\sigma}$) of a voxel $i$:

$$X_i^{\mu} = \frac{1}{|N(i)|} \sum_{j \in N_r(i)} X_j = \frac{1}{(2r+1)^3} \sum_{j \in N_r(i)} X_j \qquad \text{(Eq. 2)}$$

$$X_i^{\sigma} = \sqrt{\frac{1}{(2r+1)^3} \sum_{j \in N_r(i)} \quad X_j^2 - \left(X_i^{\mu}\right)^2} \qquad \text{(Eq. 3)}$$

The choice of the radius depends on the context and the amount of information to be aggregated. In the scope of this thesis, we choose a radius of $r = 1$, which defines a neighborhood of 26 voxels around a center voxel $i$.

### 2. Gradient Magnitude

The gradient magnitude provides complementary information regarding edge structures in the MRI data. Specifically, it offers contextual cues as to whether a voxel is located in the vicinity of a boundary between distinct tissue classes, e.g. between white and gray matter. The gradient of an MRI volume is defined as:

$$X_i^{\nabla} = \sqrt{\left(\frac{\partial X_i}{\partial x}\right)^2 + \left(\frac{\partial X_i}{\partial y}\right)^2 + \left(\frac{\partial X_i}{\partial z}\right)^2} \qquad \text{(Eq. 4)}$$

### 3. Laplacian of Gaussian

The gradient feature gives fine-details edge information. Complementary to it, we define the$LoG$, which captures coarser structures via the second spatial derivative of a Gaussian-smoothed image.

$$G(x, y, z; \sigma) = e^{-\frac{x^2+y^2+z^2}{2\sigma^2}} \qquad \text{(Eq. 5)}$$

With $\sigma$ the standard deviation, the Gaussian-scale representation of an image $X(x, y, z)$ is

$$L(x, y, z; \sigma) = G(x, y, z; \sigma) * X(x, y, z) \qquad \text{(Eq. 6)}$$

where $*$ is the convolution operator. The Laplacian operator can be applied to Eq. 6 and the $LoG$ can be expressed as

$$X^{LoG} = \nabla^2\big(G(x, y, z) * X(x, y, z)\big) = \big(\nabla^2 G(x, y, z)\big) * X(x, y, z) \qquad \text{(Eq. 7)}$$

The first term of the convolution exerts a smoothing effect and attenuates noise in the image. Furthermore, the parameter $\sigma$ regulates the degree of blur: increasing $\sigma$ enhances the smoothing effect, thereby producing an image with reduced fine detail. Consequently, the image $X^{LoG}$ will predominantly contain coarser structures. Conversely, decreasing $\sigma$ diminishes the influence of the Gaussian filter. In this case, $X^{LoG}$ will preserve finer details, but will be correspondingly more sensitive to noise. In this thesis, we used $\sigma = 2$ , which is a good balance between blurring, while keeping structure details.

#### 4. Local Binary Pattern

The $LBP$ is a texture descriptor widely employed in image analysis. The $LBP$ operator assigns a label to each voxel by thresholding the intensity values of its local neighborhood relative to the intensity of the central voxel, and then encodes the resulting binary pattern as a single integer value (Ojala et al. 1996). Let $N_r(i)$ be the set of ordered indices in a cubic neighborhood of radius $r$ around $i$.

$$N_r(i) = \{j_p\}_{p=0}^{P-1}, \quad P = (2r+1)^3 - 1 \qquad \text{(Eq. 8)}$$

where $P$ represents the number of neighbor voxels. Typically, for a 3D image with radius $r = 3$, $P = 26$. Then the $LBP$ value for a central voxel $i$ can be written as

$$X_i^{LBP} = \sum_{p=0}^{P-1} \quad s\left(X_{j_p} - X_i\right) \cdot 2^p \qquad \text{(Eq. 9)}$$

#### 5. Spatial Information

To incorporate spatial information at voxel level, we employ the Neuromorphometric mask to provide anatomical context. Specifically, this mask is used to derive two types of spatial descriptors: white matter subregions and lobar information.

White matter is partitioned into three subcompartments: periventricular (PVWM), deep (DWM), and superficial (SWM) white matter. To construct the corresponding regional masks, a global WM mask is first generated by merging the left and right WM labels, and a ventricular mask is obtained by merging the left and right ventricular labels. The PVWM mask is then defined by dilating the ventricular mask by 8 voxels and intersecting this dilated mask with the original WM mask. The SWM mask is derived by subtracting a version of the WM mask eroded by 2 voxels from the original WM mask, thereby retaining only the most peripheral WM voxels. Finally, the DWM mask corresponds to the remaining WM voxels after removing both the PVWM and SWM from the original WM mask. An example of this mask is given in Figure 1B.

For lobar parcellation, the brain is divided into four main anatomical regions: frontal, parietal, temporal, and occipital lobes. The cortical labels in the neuromorphometric atlas are grouped into these four categories, which serve as seed regions for each lobe. Each WM voxel is then assigned to the nearest lobe by computing the shortest Euclidean distance to the corresponding cortical seed regions. In Figure 1B, we observe an example of such a parcellation, where voxels are segmented into frontal, parietal, temporal, and occipital lobes.

#### 6. White Matter Intensity Energy

We conclude this section by introducing the $WMIE$ feature. Let $X^{WM}$ denote the array containing the intensities of all voxels belonging to the $WM$, as defined by the neuromorphometric mask. The $WMIE$ at voxel $i$ is then defined as

$$X_i^{WMIE} = \frac{\left(X_i - \underline{x^{WM,95}}\right)^2}{\sigma^2_{x^{WM,95}}} \qquad \text{(Eq. 10)}$$

where $x^{WM,95}$ represents the subset of white matter voxel intensities up to the 95th percentile, $\underline{x^{WM,95}}$ is the mean of this subset, and $\sigma_{x^{WM,95}}$ its standard deviation. The restriction to the 95th percentile attenuates the influence of artifacts and potential lesion-related outliers within the $WM$, thereby yielding more robust summary statistics.

### Model comparison

#### 1. Precision-Recall Curve

Model performance was evaluated using precision–recall (PR) curves, which provide a threshold-independent assessment of performance.

Precision and Recall are expressed as

$$Precision = \frac{TP}{TP + FP} \quad \text{and} \quad Recall = \frac{TP}{TP + FN}$$

The area under the curve (AUC) can then be calculated to give a summary statistic to compare across models. A higher AUC leads to better results. Another summary that can also represent the Precision-Recall (PR) curves is the Average Precision (AP). It computes a weighted mean of precisions achieved at each threshold, with the increase of recall from the previous step used as weight.

$$AP = \sum_n (R_n - R_{n-1}) P_n$$

with $R_n$ and $P_n$ the Recall and Precision at step $n$.

### 2. Dice and Modified Dice

To assess the overall quality of the segmentation, we usually use the Dice-Sørensen Coefficient (DSC).

$$DSC = \frac{2|Y \cap \hat{Y}|}{|Y| + |\hat{Y}|} = \frac{2 * TP}{2 * TP + FN + FP}$$

where $Y$ and $\hat{Y}$ are ground truth and prediction map, respectively. Although this metric is the most common for image segmentation, it is not perfect. Indeed, the Dice score only accounts for the amount of misplaced elements but does not have spatial awareness of these elements. In the context of WMH, a misplaced voxel does not have the same impact depending on its location. To tackle this issue, different dice measures can be used, such as the Weighted Dice Coefficient (WDC) (Rainio and Klén 2026).

Let $Y \subset \Omega$ denote the reference segmentation and $\hat{Y} \subset \Omega$ the predicted segmentation, where $\Omega \subset Z^3$ represents the MRI volume domain. Let $Y_0 = Y$ and $\hat{Y}_0 = \hat{Y}$ . Let $Y_i$ and $\hat{Y}_i$ denote the sets obtained by $i$ successive morphological dilations of $Y$ and $\hat{Y}$ , respectively. Let $1 > v_1 > \ldots > v_n > 0$ be a sequence of decreasing weights. We can define then the WDC as:

$$WDC = 2\frac{|Y \cap \hat{Y}| + \sum_{i=1}^{n} \quad v_i|(X_i \cap \hat{Y}_i)\backslash(Y_{i-1} \cap \hat{Y}_{i-1})|}{|Y| + |\hat{Y}| + \sum_{i=1}^{n} \quad v_i(|X_i\backslash Y_{i-1}| + |\hat{Y}_i\backslash \hat{Y}_{i-1}|)}$$

### 3. Average Symmetric Surface Distance

Another way to look at the shape of the segmentation is to compute the Average Symmetric Surface Distance (ASSD). The ASSD is the average of all the distances from points on the predicted segmentation mask to the ground truth mask, and vice versa.

$$ASSD = \frac{1}{|S(Y)| + |S(\hat{Y})|}\left(\sum_{s_A \in S(Y)} d\left(s_A, S(\hat{Y})\right) + \sum_{s_B \in S(\hat{Y})} d(s_B, S(Y))\right)$$

With $S(Y)$, $S(\hat{Y})$ the set of surface voxels of $Y$ and $\hat{Y}$, respectively, and $d(v, S(Y)) = min_{S_A \in S(Y)}||v - S_A||$ the shortest euclidean distance of an arbitrary voxel $v$ to $S(Y)$.

### 4. 95 Hausdorff Distance

In addition to the ASSD that gives an impression of the boundary of the lesion, the Hausdorff distance qualifies how far voxels are from the ground truth. Let $d(a, b)$ be the euclidean distance of a point a to an object B :

$$d(a, B) := inf_{b \in B} d(a, b)$$

The Hausdorff Distance between a ground truth $Y$ and a segmentation $\hat{Y}$ can be expressed as:

$$d_H(Y, \hat{Y}) := max\{sup_{y \in Y} d(y, \hat{Y}),\ sup_{\hat{y} \in \hat{Y}} d(Y, \hat{y})\}$$

The 95 Hausdorff Distance (HD95) can be expressed by taking the values up to the 95th quantile.

5. **Relative Volume Difference and Volumetric Similarity**

In addition to the metrics that focus on the shape and the location of the voxels, it is clinically important to have a similar lesion volume between the prediction and the manual segmentation. The Relative Volume Difference (RVD) is defined as:

$$RVD = \frac{|V - \hat{V}|}{V}$$

with $V$ and $\hat{V}$ the volume of the ground truth and the prediction, respectively.

In addition to the RVD, we define the volumetric similarity as

$$VS = \frac{|V - \hat{V}|}{|V| + |\hat{V}|}$$

**Uncertainty quantification**

The uncertainty metrics are all derived from the Shannon entropy defined by:

$$H_i = -\sum_{c=0}^{C-1} P(y_i = c|x) log_2(P(y_i = c|x))$$

1. **Voxel-scale**

$$H_i = -\sum_{c=0}^{C-1} P(y_i = c|x) log_2(P(y_i = c|x))$$

2. **Lesion-scale**

$$\underline{H_l} = \frac{1}{|L|} \sum_{i \in L} H_i$$

3. **Participant-scale**

$$\underline{H_P} = \frac{1}{|L|}\sum_{l \in L} \quad H_l$$

### Longitudinal analysis

1. **Longitudinal lesion analysis**

Longitudinal lesion analyses were performed on 12 pairs of consecutive MRI examinations. Lesion masks generated by RADAR and WHITE-Net were analysed in the common MPM space. To reduce the influence of segmentation noise, RADAR lesion masks were post-processed using three-dimensional connected-component analysis (18-connectivity), retaining only connected components comprising at least five voxels. WHITE-Net masks were analysed without post-processing.

For each pair of scans, lesion correspondence between baseline (T1) and follow-up (T2) was established by identifying overlapping connected lesions. Follow-up lesions with no overlap with any baseline lesion were classified as new. For overlapping lesions, the total baseline lesion volume was compared with the follow-up lesion volume. Lesions were classified as extending when their volume increased by more than 10%, regressing when their volume decreased by more than 10%, and stable when volume changes remained within ±10%. Lesion counts and lesion volumes were computed separately for RADAR and WHITE-Net.

Longitudinal consistency was quantified using Dice similarity coefficients computed (i) within each segmentation method between T1 and T2, and (ii) between RADAR and WHITE-Net at both T1 and T2. Lesion counts, lesion volumes and Dice coefficients were summarised using paired boxplots overlaid with individual subject values and paired trajectories.

2. **Voxelwise longitudinal analysis**

To investigate the biological characteristics of tissue uniquely identified by each segmentation method, baseline lesion voxels were assigned to one of eight mutually exclusive fate categories according to their segmentation status at follow-up. Voxels initially belonging exclusively to RADAR or exclusively to WHITE-Net were classified according to whether they subsequently became part of (i) both segmentations, (ii) the same segmentation only, (iii) the alternative segmentation only, or (iv) neither segmentation (disappeared).

For each fate category, mean baseline (T1) values were extracted from quantitative MRI maps (MTsat, R1, R2*, ICVF, ISOVF, FA, MD, orientation dispersion, proton density, g-ratio and FLAIR intensity), together with RADAR probability and voxel-wise uncertainty maps. Two additional reference regions were included: voxels belonging to the intersection of RADAR and WHITE-Net lesions at baseline and normal-appearing white matter (NAWM), defined as white matter outside the union of baseline RADAR and WHITE-Net lesions with valid quantitative MRI values.

For each longitudinal pair, mean metric values were computed within every voxel-fate category and subsequently aggregated across pairs using voxel-count-weighted averaging. For visualisation only, quantitative MRI and FLAIR values were standardised independently for each imaging metric by z-scoring the group means across fate categories, whereas probability and uncertainty maps were displayed using their original values. Results were summarised using pie charts showing the proportion of voxels assigned to each fate category and heatmaps illustrating the microstructural profiles associated with each longitudinal fate.

### 3. Biological validation

**Trail Making Test (TMT)**

Processing speed and executive function were assessed using the French version of the Trail Making Test (Reitan & Wolfson, 1985), adapted from the original instructions described by Spreen and Strauss (1991). After a practice trial, participants completed Trail A by drawing a continuous line connecting numbered circles (1-24) in ascending order as quickly as possible without lifting the pen. Following a second practice trial, participants completed Trail B by alternately connecting numbers and letters in ascending numerical and alphabetical order. When an error occurred, the examiner immediately indicated it, and participants were required to correct it before continuing. Completion time, including the time required for error correction, was recorded for both parts. The same standardized forms (used at the CHUV Memory Clinic) were administered at both assessment waves. Higher completion times indicate poorer performance, with TMT-A primarily reflecting processing speed and TMT-B primarily reflecting executive function, particularly cognitive flexibility and set-shifting.

**Analyses**

Biological validation analyses compared white matter hyperintensity (WMH)-derived MRI metrics obtained from RADAR-WMH and WHITE-Net segmentations in relation to age, systolic blood pressure (SBP), and cognitive performance (TMT-A and TMT-B). Mean MRI metrics were extracted within each WMH segmentation, including WMH volume, MTsat, R1, R2*, ICVF, FA, orientation dispersion (OD), g-ratio, ISOVF, MD, and proton density (PD*). WMH volume was log-transformed prior to analysis. For each clinical outcome (age, SBP, TMT-A, and TMT-B), a common maximum-N dataset containing complete observations for all MRI metrics was constructed to ensure identical participant samples across comparisons.

***Comparison of MRI metrics between segmentation methods***

Differences in MRI metric distributions between RADAR-WMH and WHITE-Net were assessed using paired two-sided *t*-tests for each metric. Resulting *P*-values were corrected for multiple comparisons using the Benjamini–Hochberg false discovery rate (FDR) procedure.

***Biological sensitivity analyses***

To determine whether MRI metrics derived from each segmentation exhibited differential sensitivity to clinical variables, standardized linear regression models were fitted separately for each MRI metric and clinical outcome. MRI metrics and continuous clinical variables were z-scored before analysis.

The following model was fitted:

$$MRI\ metric_z \sim Clinical\ variable_z \times Method + Sex + Education + C(Subject)$$

The interaction term tested whether associations differed significantly between segmentation methods. Subject was included as a repeated factor to account for paired RADAR-WMH and WHITE-Net measurements within individuals. Age was additionally included as a covariate in supplementary age-adjusted SBP analyses. SBP was measured in mmHg as the average of the second and third out of three measurements for each timepoint. Resulting p-values were corrected for multiple comparisons using FDR correction.

Predictive value was evaluated using repeated 5-fold cross-validated ridge regression (50 repetitions). Baseline models included demographic covariates (sex and education), and MRI metrics from each segmentation method were added separately.

Baseline model:

$$Clinical\ variable \sim Sex + Education$$

RADAR-WMH model:

$$Clinical\ variable \sim RADAR_{MRI\ metric} + Sex + Education$$

WHITE-Net model:

$$Clinical\ variable \sim WHITE\ Net_{MRI\ metric} + Sex + Education$$

Predictive improvement was quantified as the increase in out-of-sample explained variance relative to the baseline model:

$$\Delta R^2 = R^2_{RADAR-WMH\ model} - R^2_{Baseline\ model}$$

Differences in $\Delta R^2$ between RADAR-WMH and WHITE-Net were assessed using paired permutation testing (10,000 permutations) with FDR correction.
